# Early interventions for first onset of symptoms of mental health conditions: an umbrella review of systematic reviews

**DOI:** 10.1101/2025.05.08.25323765

**Authors:** Jasmine Lee, Phoebe Barnett, Jialin Yang, Rebecca Appleton, Brynmor Lloyd-Evans, Jane Hahn, Nathalie Rich, Emma Francis, Lizzie Mitchell, Eva Driskell, Alina Unkelbach, Sonia Johnson

## Abstract

**Background:** Early intervention following mental health symptom onset has great potential in reducing long-term burden on individuals, families and friends, and society. The main focus in service development and research has been on early intervention in psychosis, but advances have also been made for other mental health difficulties such as eating disorders, anxiety and depression. We aimed to take stock of the available evidence regarding effectiveness, implementation, and experiences of care for early intervention approaches through a systematic umbrella review.

**Methods:** We included systematic reviews of complex early intervention strategies including more than one component, for mental health conditions with typical onset in young people under 25. We searched 4 databases (January 2019-April 2024) and synthesised results from eligible reviews using a narrative approach. Quality was assessed using AMSTAR 2. We excluded reviews on At Risk Mental States for psychosis as this is an extensive literature that has been the sole focus of umbrella reviews.

**Results:** Sixteen reviews were included, with ten covering early intervention for psychosis, three for eating disorders, one for bipolar disorder and two for transdiagnostic early intervention approaches. Reviews of early intervention for psychosis suggest that intensive approaches can improve recovery rates following first presentation to services, although the success of initiatives aimed at reducing duration of untreated symptoms is less consistent. We found little systematically synthesised evidence of good quality regarding other diagnoses, although some early indications of success with eating disorders were described. Stigma and lack of knowledge or support act as barriers to rapid access, while insufficient service resources and staffing were barriers to effective intervention delivery.

**Conclusions:** Despite its high importance in reducing the global burden of mental ill-health, evidence on how to intervene early following symptom onset remains limited (as assessed via systematic reviews), especially for conditions other than psychosis. For psychosis, some approaches now warrant attention to widespread implementation. Innovative approaches for eating disorders have been developed, but there is still a pressing need for treatments supported by substantial and robust trials. Further systematic reviews would be desirable for conditions including depression and anxiety, bipolar disorder, and “personality disorder”.

## Introduction

Early intervention services aim to identify and treat mental health difficulties as early as possible and thus improve prognosis. Research suggests that around two-thirds of mental health problems have their onset between ages 14-24 (Girolamo et al., 2012; Kessler et al., 2005) and in 2019, mental health conditions were the leading cause of disability among young people in Europe (Castelpietra et al., 2022). This highlights a pressing need to improve intervention efforts and research focusing on this illness stage (Colizzi et al., 2020).

Preventative approaches to mental health problems include strategies for the entire population (universal prevention), for those at greater risk of developing problems (selective prevention), and for those presenting with early signs (indicated prevention) (Patricia J. Mrazek & Haggerty, 1994). Indicated prevention typically involves early intervention in primary and community mental health care, targeting individuals who present with early symptoms of a condition. The current review focuses on this, and also on secondary prevention to reduce the impact of illness on people who are still in the early stages of a mental health problem, but now reach a diagnostic threshold for a formal diagnosis (Callaly, 2014). Indicated and secondary prevention are often mingled within the same service for people with early symptoms that may be just above or below a diagnostic threshold (Fusar-Poli et al., 2021), and are thus discussed together in this review.

Traditionally, mental health care has adopted a more reactive as opposed to preventative approach (McGorry & Mei, 2018), however in recent decades early intervention services, which aim to react more quickly, have come to the fore for psychotic disorders, and these models have been shown to improve outcomes and reduce costs (McGorry, 2015; Ricciardi et al., 2008). As well as reducing the long-term impact of mental ill health, early intervention has the potential to improve physical health outcomes such as rates of cardiometabolic disease (Shiers & Lester, 2014). Other reasons to intervene early in mental health conditions include reducing disruption of relationships with family, friends and wider community, maintaining pathways through education and employment (5), and reducing the likelihood of serious incidents occurring while mental health problems remain untreated (Callaly, 2014; Shiers & Lester, 2014).

However, there remain several challenges that hinder broader implementation of early intervention approaches. In the UK, many specialist mental health services, such as Children and Young People’s Mental Health Services (CYPMHS, formerly CAMHS) and Eating Disorder services, have high clinical thresholds including severe mental health symptoms and impaired daily functioning for receipt of support – often at ‘crisis point’ (Holding et al., 2022; Marlowe, 2014). Minoritised groups, people from lower-income backgrounds, disabled people and older (aged 65 or over) people, are also more likely to face delays to initial treatment, have poorer experiences when receiving care, and have reduced access to services (Bansal et al., 2022; Department of Health and Social Care, 2023; Kapadia et al., 2022; Office for National Statistics, 2022). Such barriers may contribute to the exacerbation of symptoms which may be prevented if support was offered earlier. Financial and staffing constraints have further limited availability and effectiveness of early intervention efforts.

Although funding for mental healthcare has increased in recent years, with the UK spending £12 billion on mental health services in England in 2021/2022, this financial increase is not enough to keep up with the increasing demand (Gilburt & Mallorie, 2024; Islam et al., 2023). Furthermore, the potential of early intervention for first-episode presentations of common mental health problems like depression, anxiety and eating disorders has received less attention than approaches for individuals with early signs of severe mental illness (Colizzi et al., 2020). This lack of clear models that are underpinned by theory and evidence is likely to impede early intervention efforts (Hetrick et al., 2017; Settipani et al., 2019; Yung, 2016).

Research-based consensus on the best approaches to supporting the full range of mental health problems experienced by those presenting to community-based early intervention services is thus still limited. Our aim in this umbrella review was to take stock of evidence currently available to inform service development and to identify gaps, by addressing the following research questions:

1. What evidence is available from systematic reviews on the effectiveness of early intervention models in the community for people with early symptoms of mental health conditions?
2. What are the facilitators and barriers to these models being implemented as intended and achieving their aims?
3. What are service users’, carers’, and staff’s experiences of these services?

## Methods

This umbrella review was conducted by the NIHR Policy Research Unit in Mental Health, based across University College London and King’s College London, which presents independent research to inform government and NHS policy in England. It was conducted according to Cochrane guidelines (Pollock et al., 2022) and written according to Preferred Reporting Items for Systematic Reviews and Meta-Analyses (PRISMA) guidelines [**see Appendix 1 for PRISMA checklist]**. The protocol was prospectively registered on PROSPERO (registration number: <u>CRD42024541486</u>).

The protocol was followed apart from the following deviations:

1. Although not explicitly specified in the protocol, we included reviews of carer perspectives;
2. We did not exclude studies which only reported acceptability if they met all other inclusion criteria.

### Search strategy

We searched four electronic databases: MEDLINE via OVID; PsycINFO via OVID; Embase via OVID and The Cochrane Database of Systematic Reviews (CDSR) for relevant systematic reviews published within the last five years (between January 2019 and April 2024). The search strategy combined terms for mental health disorders, early intervention, and systematic reviews [**see Appendix 2 for full search strategy**]. There was no date limit for the primary papers included in reviews and no language restrictions were imposed on the search. Backward citation searches for relevant systematic reviews within the date limits were also conducted.

### Eligibility criteria

We included reviews meeting the following criteria:

#### Population

*Included:* Populations aged <65, experiencing early symptoms of mental health conditions with a typical peak onset between 11-25, principally depression, anxiety disorders, psychotic disorders, trauma-related conditions, and difficulties resulting in a personality disorder diagnosis.

*Excluded:* Reviews focusing specifically on populations with neurodevelopmental conditions, dementia, or substance use (without co-occurring mental health symptoms), or reviews of services for specific occupational or physical comorbidity sub-groups.

#### Intervention

*Included:* Early intervention services or approaches for populations experiencing first onset of mental health symptoms which were designed to increase the speed or ease of access to care, or provide targeted interventions to improve outcomes following the onset of symptoms.

We included models that were intended as improvements on usual care for each condition, and that met criteria for complex interventions (Thomas et al., 2019). This was defined as:

1. Care delivered by more than one person, or
2. Care consisting of multiple components (e.g. psychotherapy AND peer support), or
3. Interactions between components or contexts of an intervention (e.g. next phase of care administered after a threshold is reached).

*Excluded:* Reviews of universal or selective prevention, or treatment aimed at recurrent mental health conditions were not eligible. We also decided to exclude systematic reviews on identification and management of individuals who are at clinically high risk for psychosis (CHR-P), as there is an extensive and complex body of literature that has by itself been the subject of umbrella reviews (Andreou et al., 2023; Frearson et al., 2025; Poletti et al., 2024).

#### Context

*Included:* Community based (mainly outside of hospital care or residential services), or services that work with people during acute admissions as well as in the community.

*Excluded:* Reviews of online-only interventions were not eligible, although interventions provided across a range of formats including online provision were included.

#### Outcome

*Included:* Reviews were required to report at least one of:

- The effectiveness of early intervention services/approaches (duration of untreated illness or change in symptom severity, quality of life, social functioning, or goal-based outcomes such as employment).
- Implementation outcomes, and facilitators and barriers to implementation.
- Experiences of service users, carers or staff.

*Excluded:* Reviews reporting only cost-effectiveness outcomes or barriers to more general help-seeking for mental health support were excluded.

#### Study designs

*Included:* Published peer-reviewed systematic reviews with or without meta-analyses, realist reviews, rapid reviews, scoping reviews, or qualitative meta-syntheses. We defined systematic reviews as those that searched at least three different bibliographic databases and used systematic methods to minimise bias. Quantitative systematic reviews were also required to have conducted a quality appraisal of included studies – this did not apply to qualitative reviews or scoping reviews as quality appraisal of studies for these is yet to become standard practice (Peters et al., 2021).

*Excluded:* Non-systematic, narrative reviews, protocols of reviews, and umbrella reviews were not eligible.

### Screening

After de-duplication, 25% of titles and abstracts were dual-screened independently by two reviewers (JL, JY) in Covidence (*Covidence Systematic Review Software*, n.d.) to ensure consistencies of application of eligibility criteria. The remaining 75% were screened by one reviewer. At the full text screening stage, 100% of reviews were independently double screened by the review team (JL, JY, JH, NR, PB), with discrepancies resolved through team discussion and consultation with a senior member of the team. Reasons for exclusion of all reviews assessed at full text were noted.

### Data extraction

Data extraction was conducted in Microsoft Excel after piloting the extraction form on 10% of included reviews and making any necessary amendments. Data for each included paper were extracted in duplicate by two of four independent members of the review team (JL, JY, JH, NR, PB), with discrepancies identified and resolved. The data extracted included information about reviews (e.g. review type, objectives, number of included studies), primary studies (e.g. date range, study designs), search strategies (e.g. databases, inclusion/exclusion criteria), participant details (e.g. gender, age, mental health condition), additional information (e.g. quality appraisal, conclusions and limitations), and reported outcomes of the reviews (e.g. types of services, effectiveness, implementation facilitators/barriers, and service user experiences).

### Quality appraisal

Quality appraisal was also conducted in duplicate by two of the four review team members (JL, JY, JH, NR) using the AMSTAR 2 Checklist (A MeaSurement Tool to Assess systematic Reviews) (Shea et al., 2017). Given the broad range of review types included in this umbrella review, such as scoping reviews and qualitative meta-syntheses, we adapted AMSTAR 2 for scoping reviews and qualitative reviews following the method used by Cooper et al. (Cooper et al., 2024) [**see Appendix 3]**. The review team independently assessed reviews blind with conflicts resolved through discussions between two authors (JL and JY). Review quality was assessed according to guidance by Shea and colleagues (Shea et al., 2017) by focusing on the number of critical or non-critical weaknesses. These were also adapted as outlined in Appendix 3.

Reviews without meta-analyses were not assessed on meta-analytical methods and risk of bias of individual studies in meta-analyses, nor publication bias. Rapid reviews were scored on the same criteria as for systematic reviews, following Cochrane guidance (i.e. including the critical domain of risk of bias assessment) (Garritty et al., 2024).

### Evidence synthesis

We synthesised data for each of our review questions using a narrative approach (Rodgers et al., 2009), grouping reviews by their population focus (mental health condition) and subsequently by the characteristics of early interventions they included. Effectiveness outcomes were narratively synthesised as there was not enough homogenous meta-analytic data to be combined meaningfully, however meta-analytic effect sizes were reported if included in the original review. Where only some models of support met our inclusion criteria, we provide information on the studies included in reviews as well as specifically those meeting our inclusion criteria and for which we report outcomes of here.

### Lived experience researcher involvement

Three lived experience researchers (experts by personal experience of mental health difficulties) were part of the research team and involved at various stages throughout the project, including attending regular team meetings, reviewing the systematic review protocol, contributing to synthesis of results and write-up.

## Results

### Study selection

The search identified 2907 references, of which 112 potentially relevant full-text articles were assessed for eligibility, following which 96 were excluded [**see Appendix 4 for list with reasons for exclusion]**. Backward citation searching did not identify additional reviews meeting inclusion criteria. Sixteen reviews met eligibility criteria and were included. Figure 1 provides further information on the full search and screening procedure.

**Figure 1:**
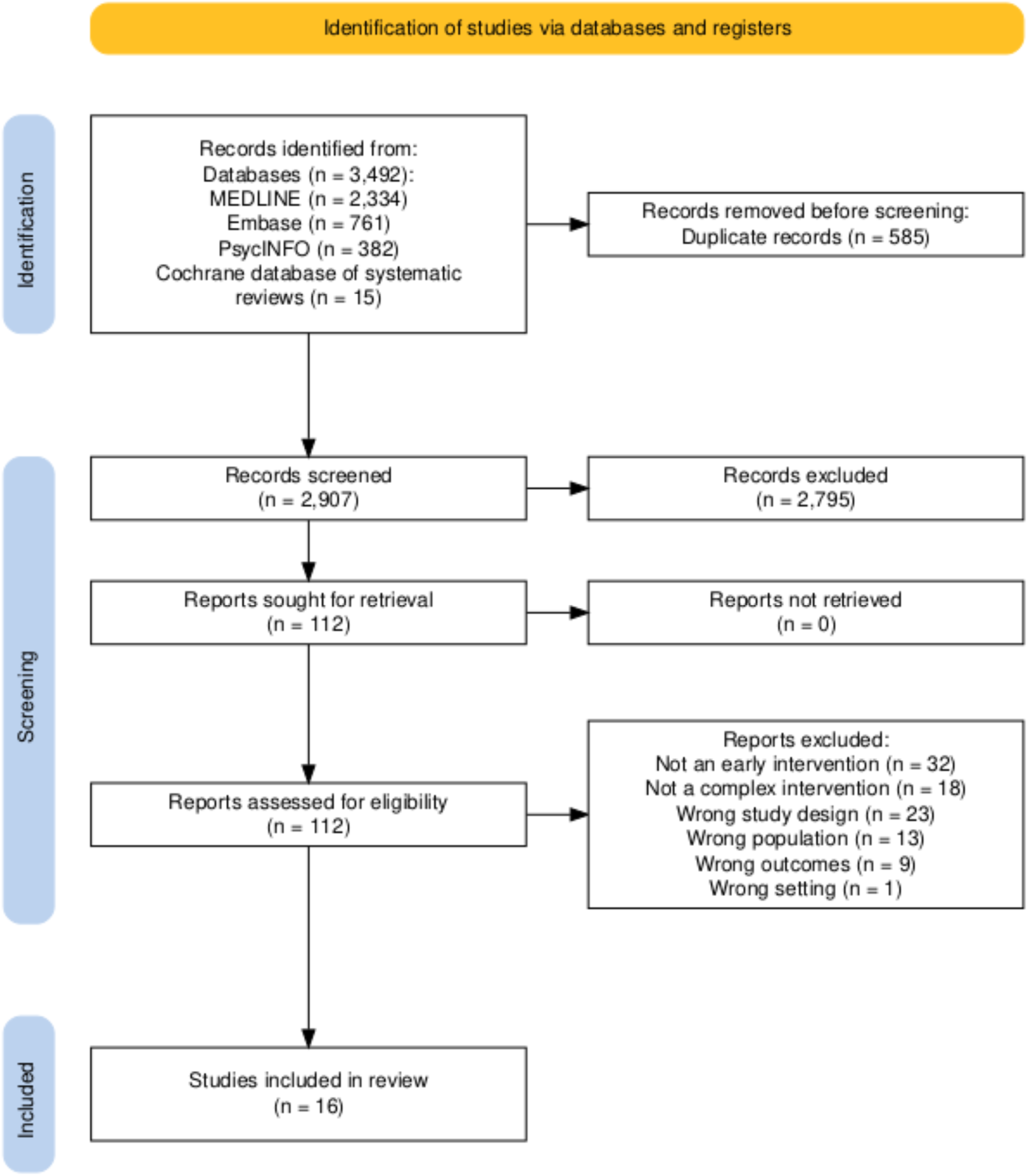
PRISMA flow diagram.

### Quality of included reviews

According to modified AMSTAR 2 ratings, 2/16 reviews were of critically low quality, 4/16 were of low quality, 8/16 were of moderate quality, and 2/16 were of high quality.

The most common critical limitations included: not accounting for risk of bias in individual studies when interpreting and discussing results of the review (3/9 reviews which were not qualitative or scoping designs), and not registering a protocol prior to the conduct of the review (3/16 reviews).

### Study characteristics

Of the 16 included reviews, three were quantitative systematic reviews with narrative syntheses (Farooq et al., 2024; Murden et al., 2024; Ratheesh et al., 2023), three were scoping reviews (Aceituno et al., 2021; Jongen et al., 2023; Settipani et al., 2019), two were quantitative systematic reviews with meta-analyses (Puntis et al., 2020; Salazar de Pablo et al., 2024), two were rapid reviews (Koreshe et al., 2023; Pehlivan et al., 2022), two were systematic reviews and narrative syntheses of facilitators and barriers (including mixed methods primary studies (O’Connell et al., 2021; Tiller et al., 2023)), two were thematic meta-syntheses (Causier et al., 2024; Loughlin et al., 2020), one was a systematic review and components network meta-analysis (Williams et al., 2024), and one was a mixed-methods systematic review to inform a health technology assessment (HTA) (Hamson et al., 2023).

Ten reviews focused on early interventions for first-episode psychosis (FEP; (Aceituno et al., 2021; Causier et al., 2024; Farooq et al., 2024; Loughlin et al., 2020; Murden et al., 2024; O’Connell et al., 2021; Puntis et al., 2020; Salazar de Pablo et al., 2024; Tiller et al., 2023; Williams et al., 2024)). Three focused on early interventions for eating disorders (Hamson et al., 2023; Koreshe et al., 2023; Pehlivan et al., 2022). One explored early interventions for bipolar disorder, although only one of the included studies examined an intervention considered complex and is thus described in this review (Ratheesh et al., 2023). Two reviews included transdiagnostic early intervention models for a variety of mental health problems, specifically in young people (Jongen et al., 2023; Settipani et al., 2019), although one had a broader intervention focus on health pathways for indigenous youth, including one primary study meeting our criteria. We did not find reviews of early intervention approaches for specific common mental health problems such as depression, nor for symptoms associated with a personality disorder diagnosis.

Reviews covered studies primarily conducted in high or middle-income countries (n=14), such as the UK, USA, Canada, and Australia. One review (Farooq et al., 2024) focused on low- or middle-income countries (LMICs), including studies conducted in India, Iran, Nigeria, Nepal, Tunisia, and Uganda. Another review (Aceituno et al., 2021) focused on Latin American settings, specifically in Argentina, Brazil, Chile, and Mexico. The sample size of primary studies included in reviews ranged from 5 to 36,309, with the total range of ages included in samples ranging from 10 to 60 years.

Eleven reviews reported on the effectiveness of the intervention in improving outcome measures (Aceituno et al., 2021; Farooq et al., 2024; Hamson et al., 2023; Jongen et al., 2023; Koreshe et al., 2023; Pehlivan et al., 2022; Puntis et al., 2020; Ratheesh et al., 2023; Salazar de Pablo et al., 2024; Settipani et al., 2019; Williams et al., 2024) and six on reducing duration of untreated illness (Aceituno et al., 2021; Hamson et al., 2023; Koreshe et al., 2023; Murden et al., 2024; Pehlivan et al., 2022; Salazar de Pablo et al., 2024). Eight reported outcomes relating to implementation, including barriers and facilitators to successful services and patient access (Aceituno et al., 2021; Causier et al., 2024; Farooq et al., 2024; Jongen et al., 2023; O’Connell et al., 2021; Pehlivan et al., 2022; Settipani et al., 2019; Tiller et al., 2023). Three reviews reported outcomes relating to experiences of care (Loughlin et al., 2020; Puntis et al., 2020; Salazar de Pablo et al., 2024).

We explored the extent of overlap in included primary studies to ascertain whether results of some studies may bias overall conclusions. Only two primary studies were included in three reviews, while 26 studies were included in two reviews [**see Appendix 5 for overlapping studies]**.

Only 4 out of 16 reviews (Causier et al., 2024; Hamson et al., 2023; Koreshe et al., 2023; Pehlivan et al., 2022) involved lived experience researchers (LERs) in the study design process. Of these 4 reviews, 3 focused on early intervention for eating disorders, and one for psychosis. Further characteristics of each study are summarised in the following sections.

### Data synthesis

#### Early interventions for psychosis

Ten reviews synthesised research on early intervention approaches for psychosis. One review synthesised evidence for strategies designed to reduce the Duration of Untreated Psychosis (DUP) and improve pathways to care (Murden et al., 2024), and two qualitative meta-syntheses described structural barriers that deter patients and carers from seeking help from early intervention models (Causier et al., 2024; Tiller et al., 2023). Three reviews synthesised findings relating to outcomes of early intervention models with a main aim of improving prognosis for patients experiencing First-Episode Psychosis (FEP) once they have presented to services (Farooq et al., 2024; Puntis et al., 2020; Williams et al., 2024), and we also included two qualitative syntheses of facilitators to successful implementation (O’Connell et al., 2021) and experiences of initial engagement with these service models (Loughlin et al., 2020). Two reviews (Aceituno et al., 2021; Salazar de Pablo et al., 2024) included interventions across both primary aims of reducing DUP and improving prognosis. Tables 1 and 2 describe individual review characteristics and outcomes for strategies to reduce DUP and improve pathways to care, and strategies to improve prognosis, respectively.

**Table 1:**
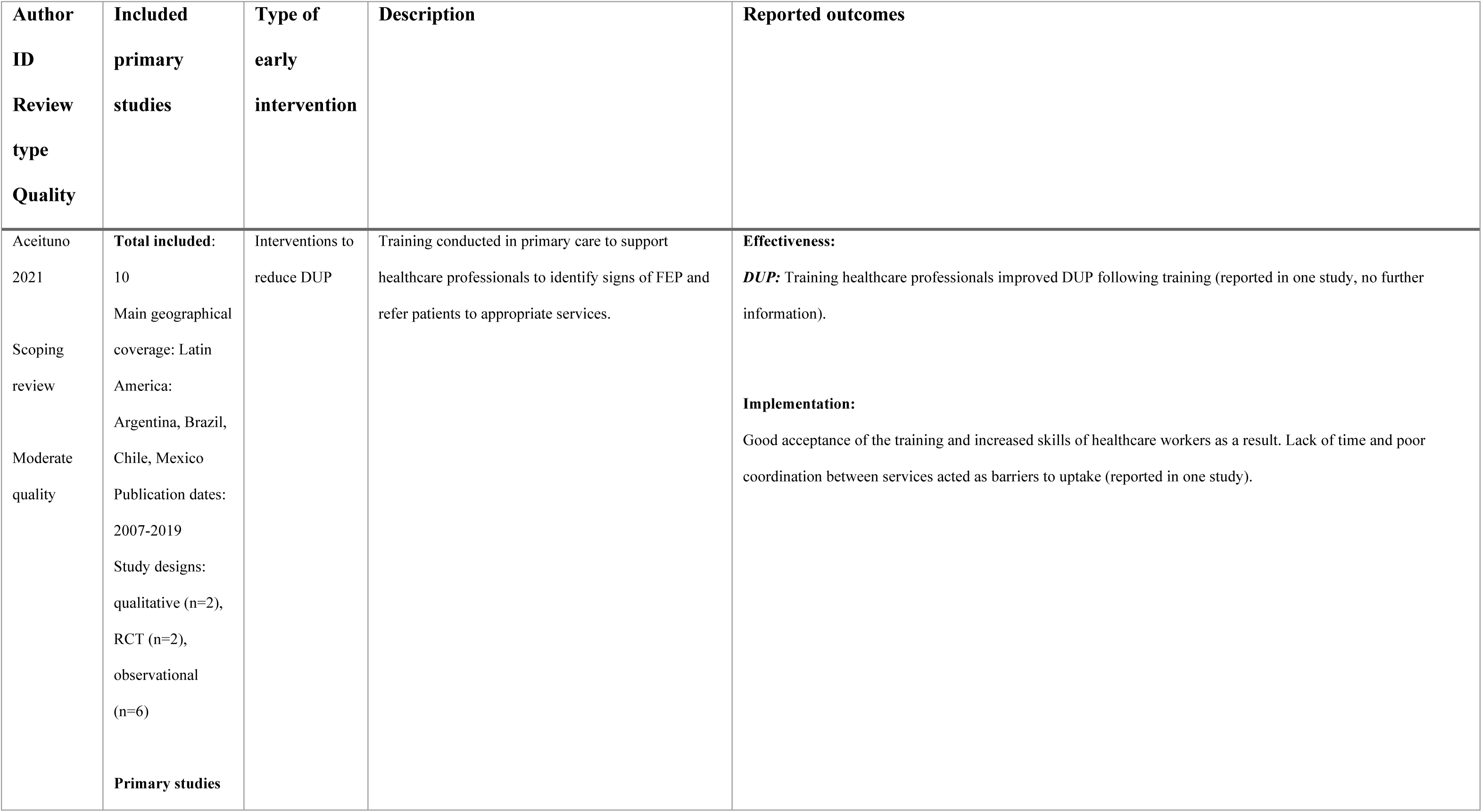
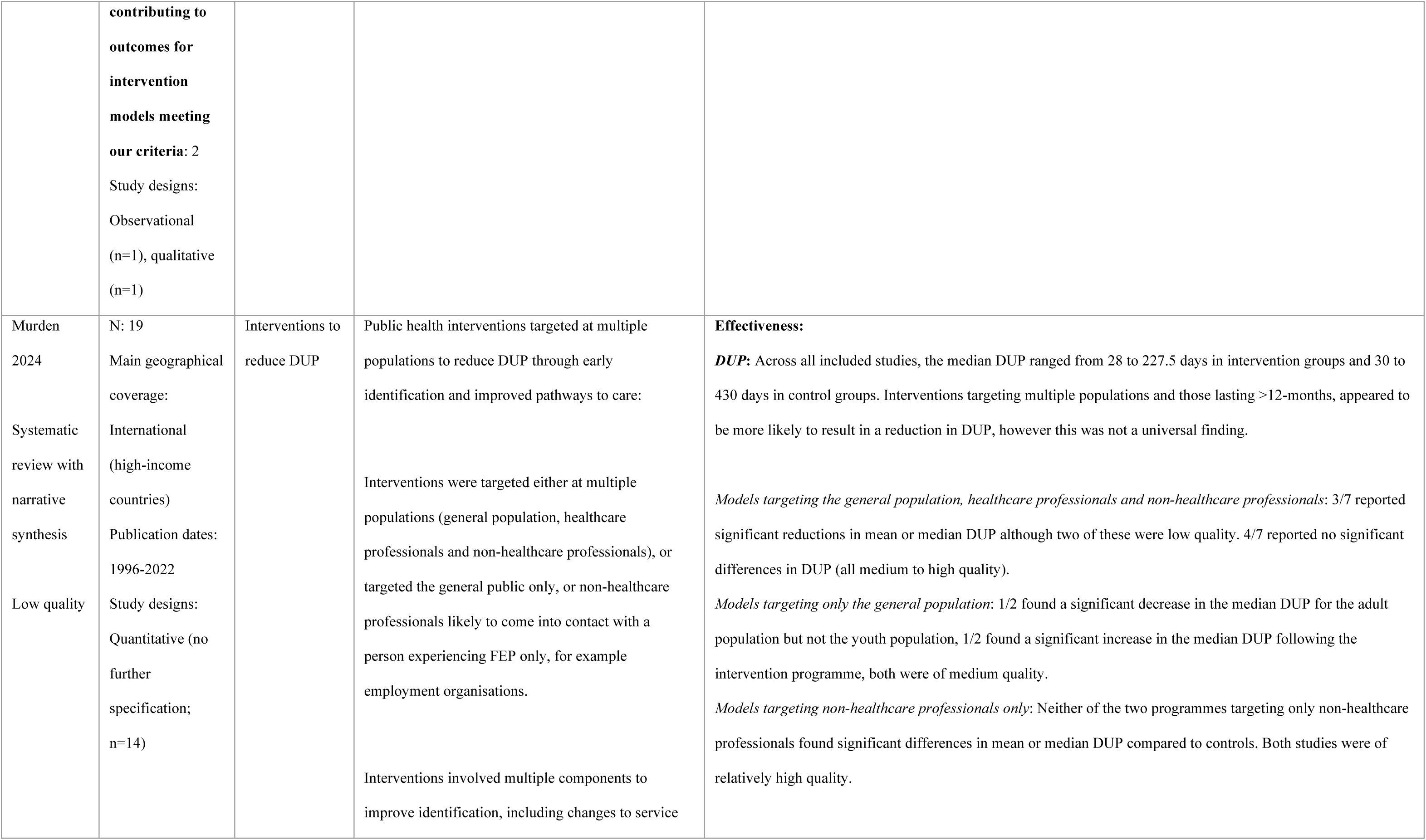
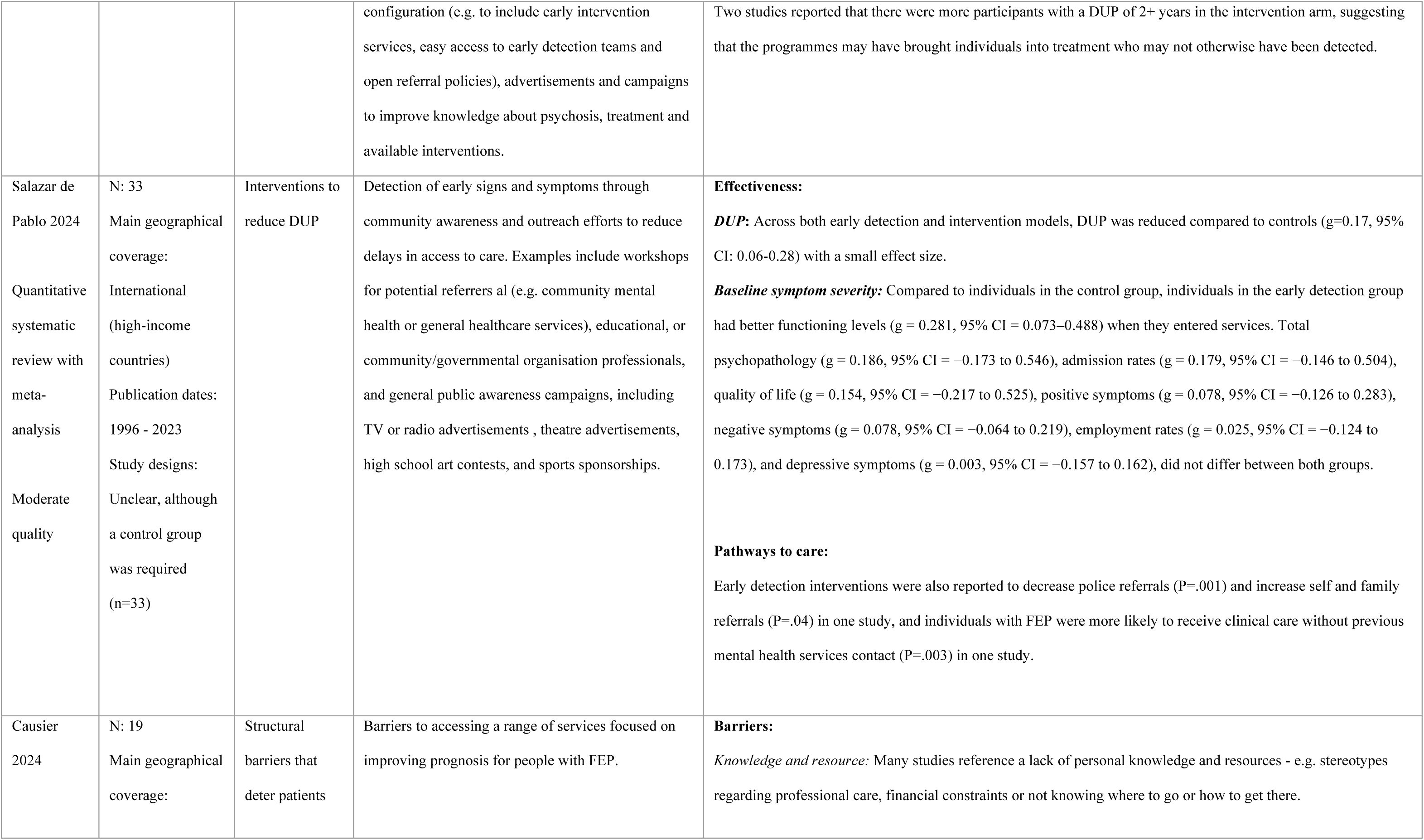
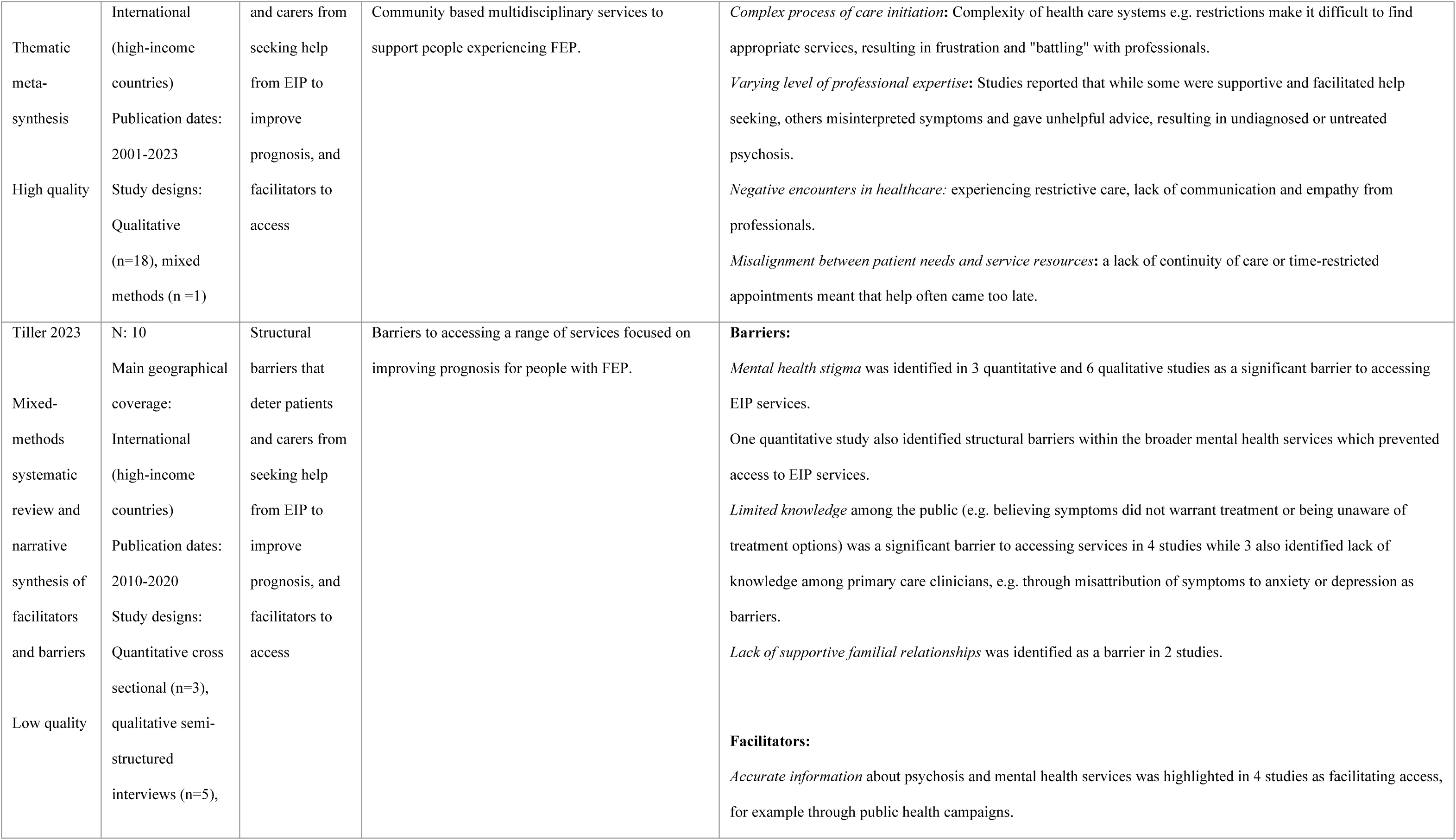
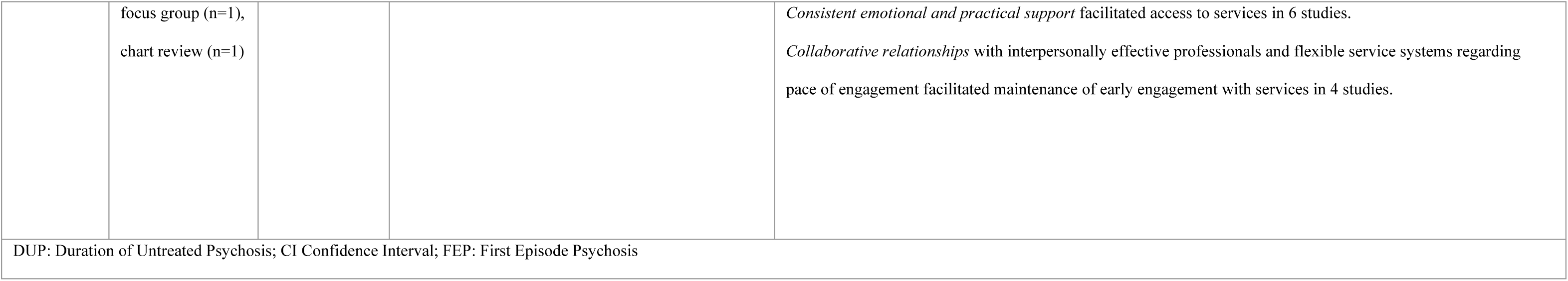
Reviews of early intervention models aimed at reducing DUP and improving experiences of pathways to care.

**Table 2:**
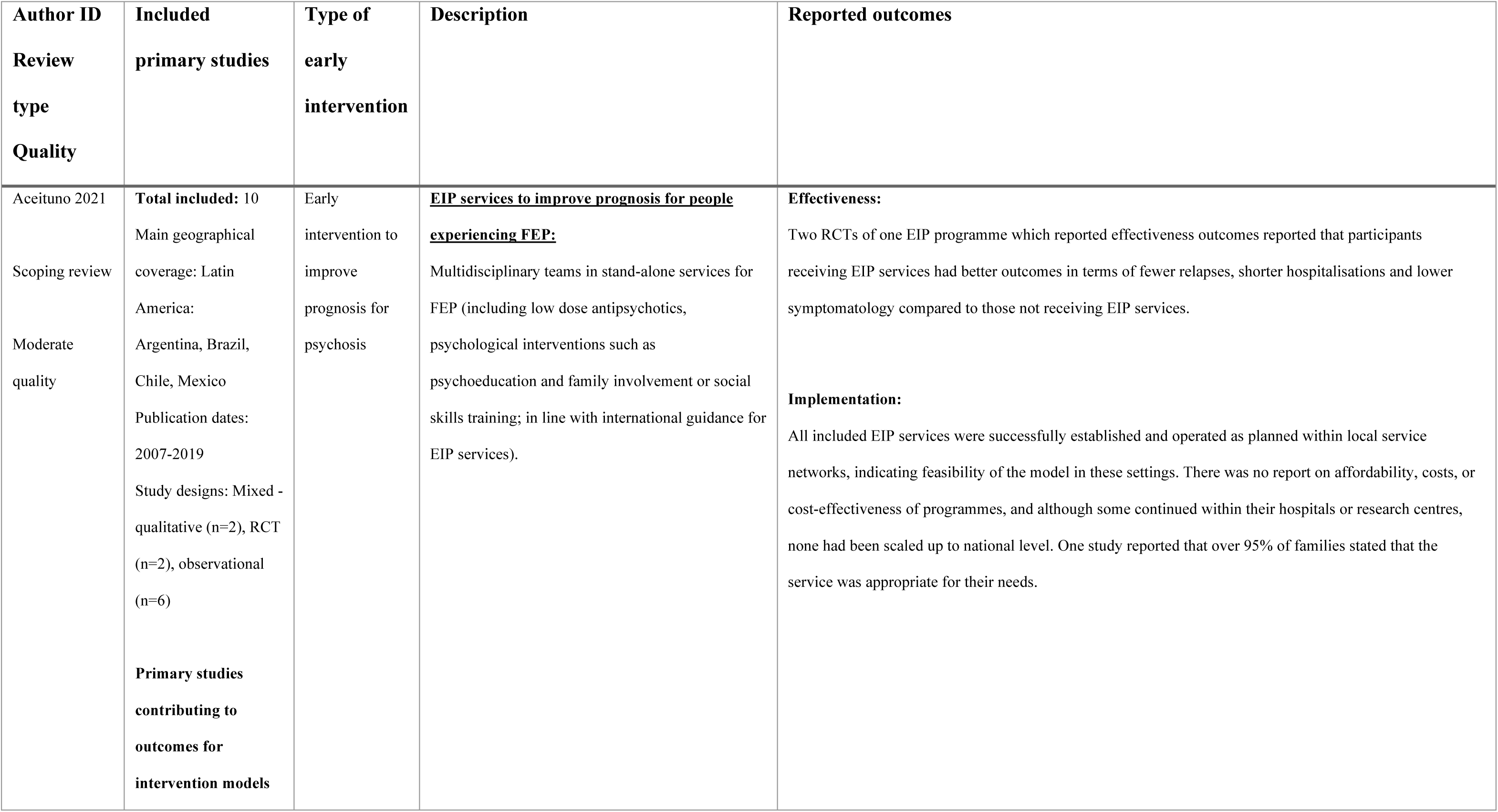
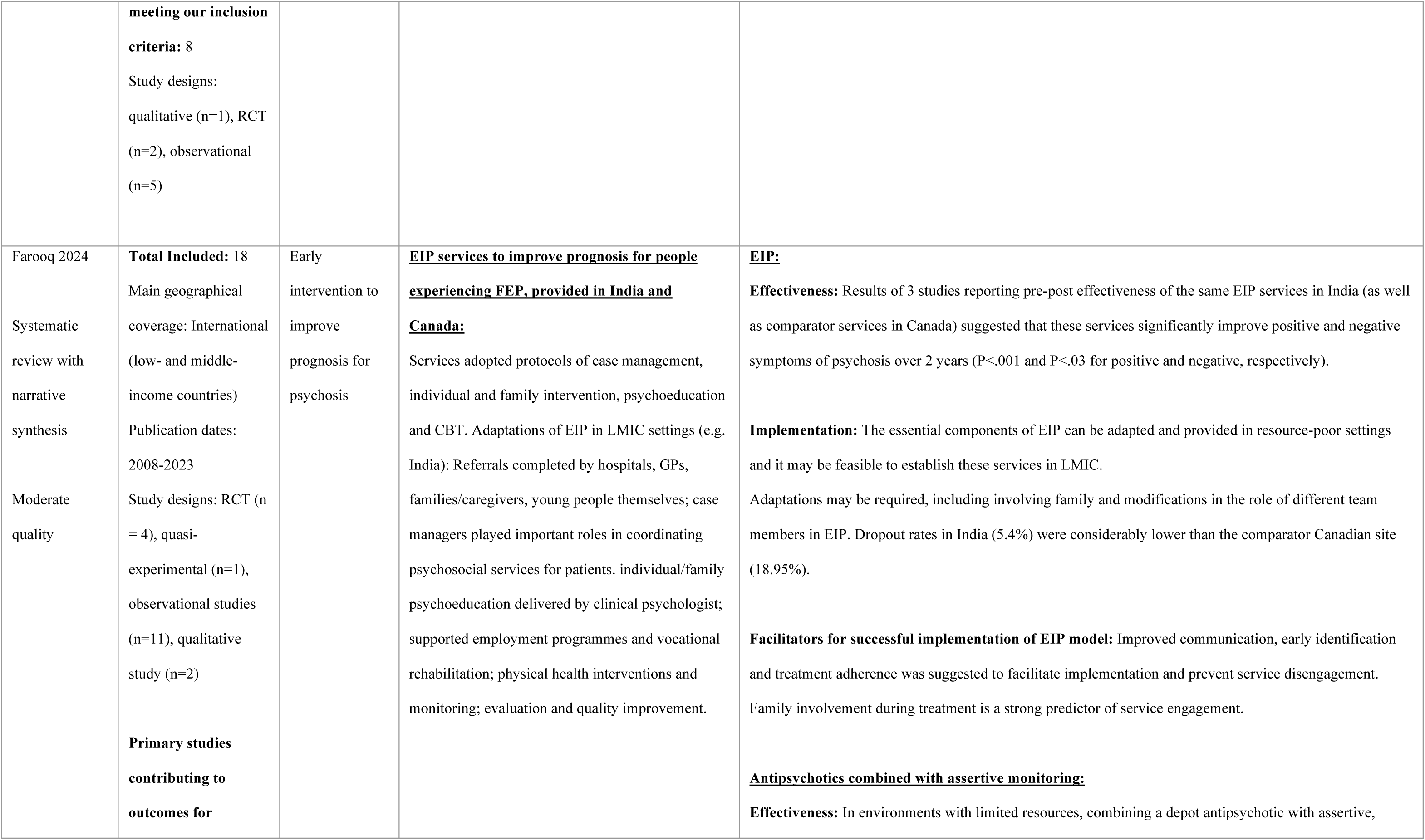
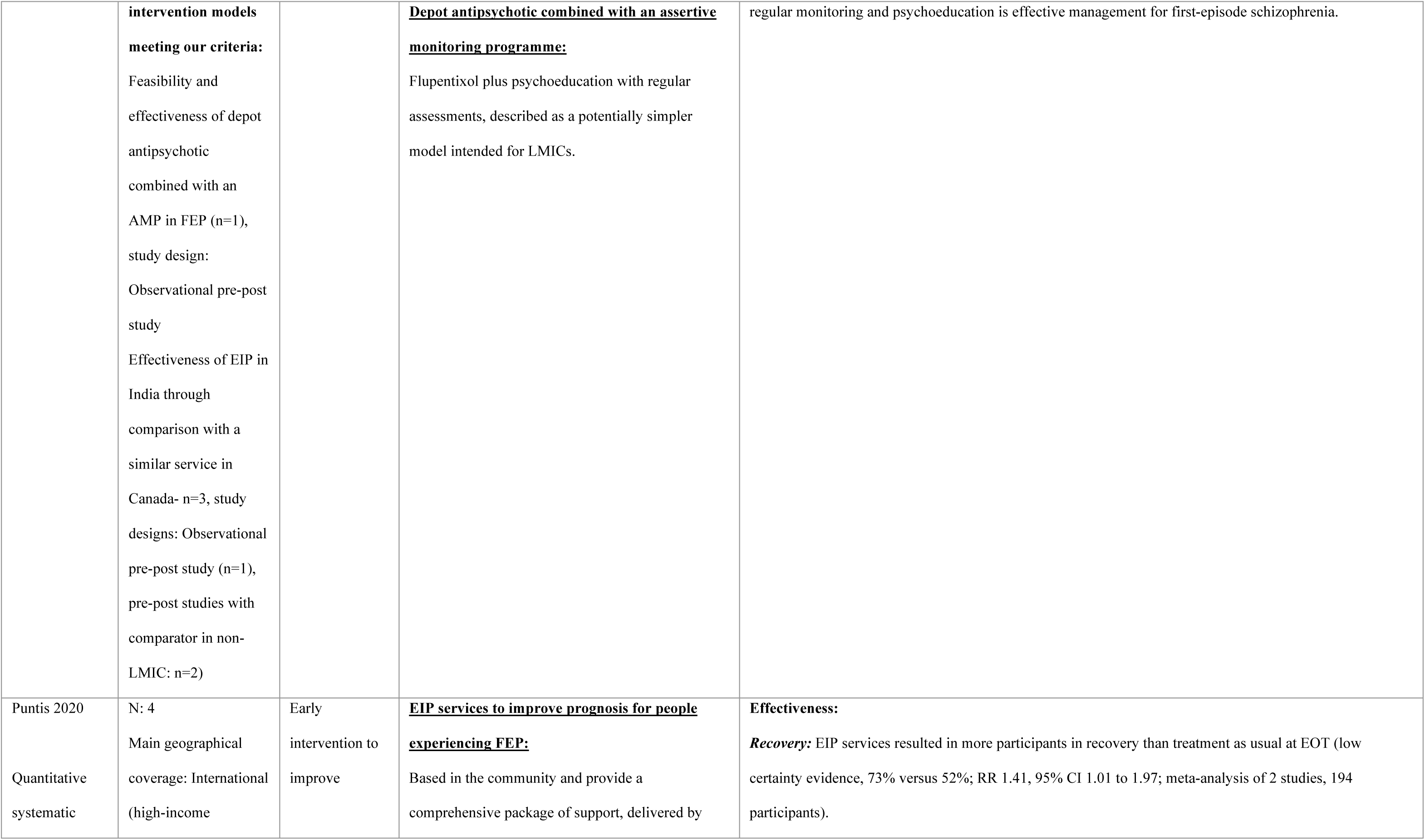
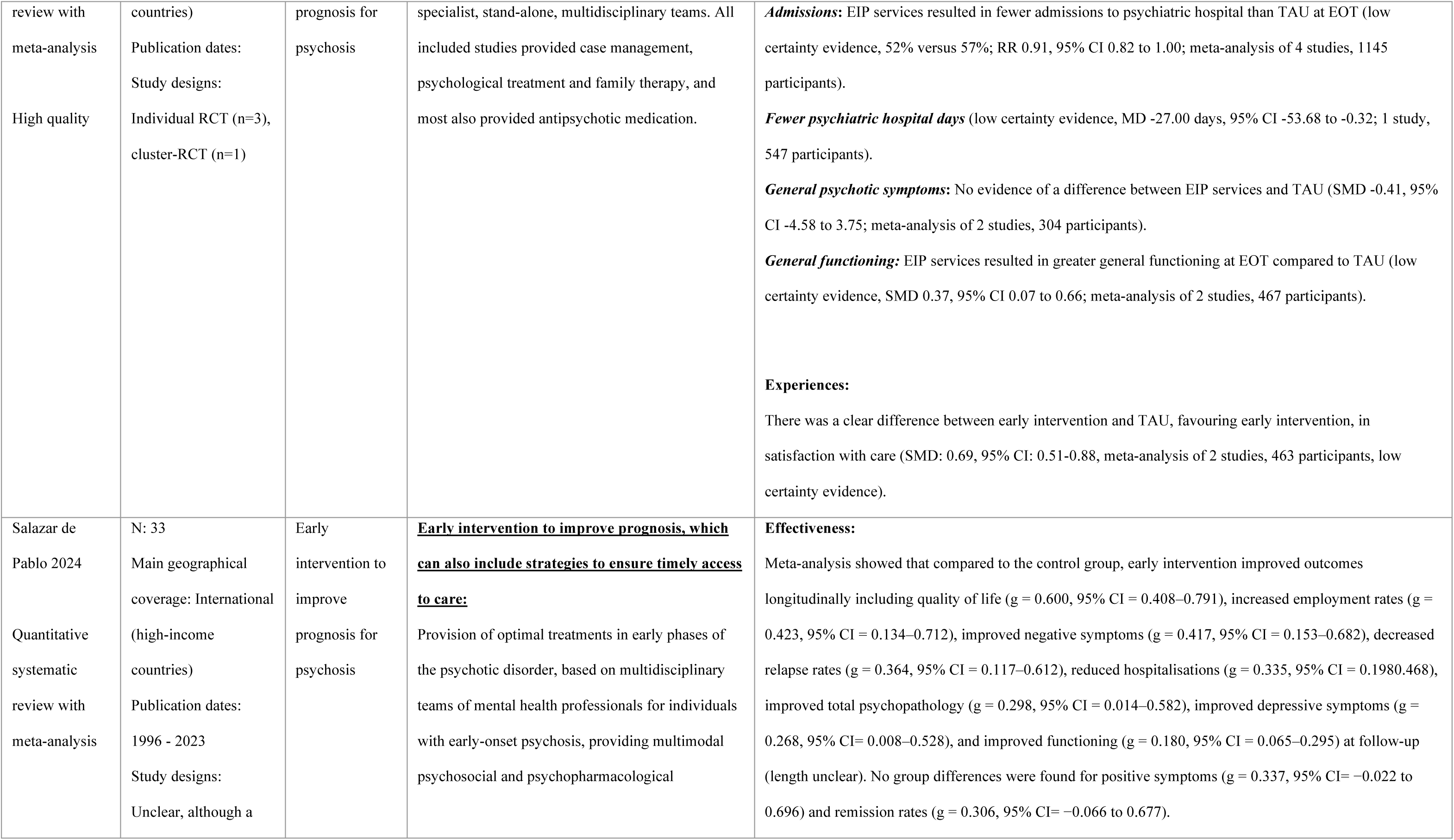
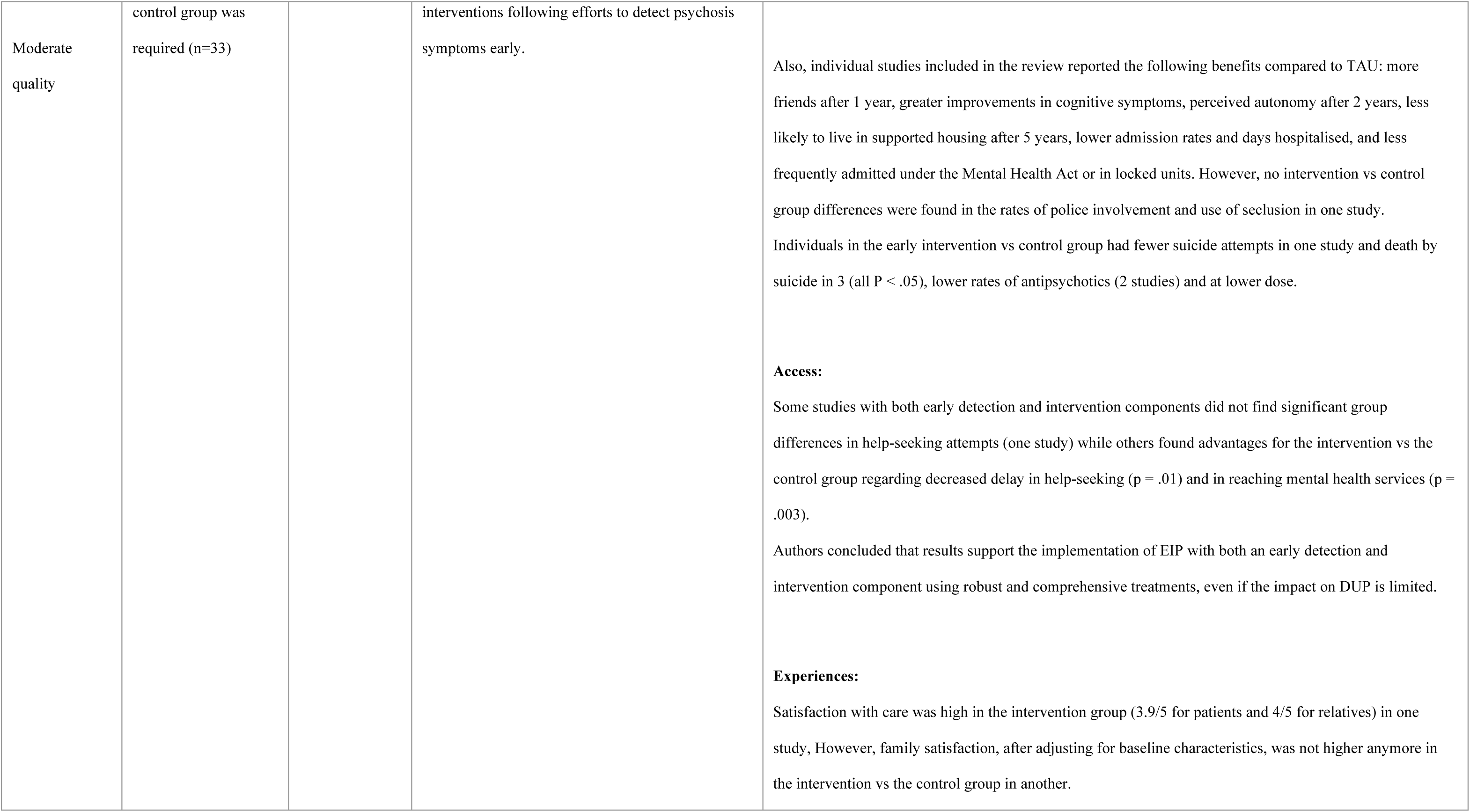
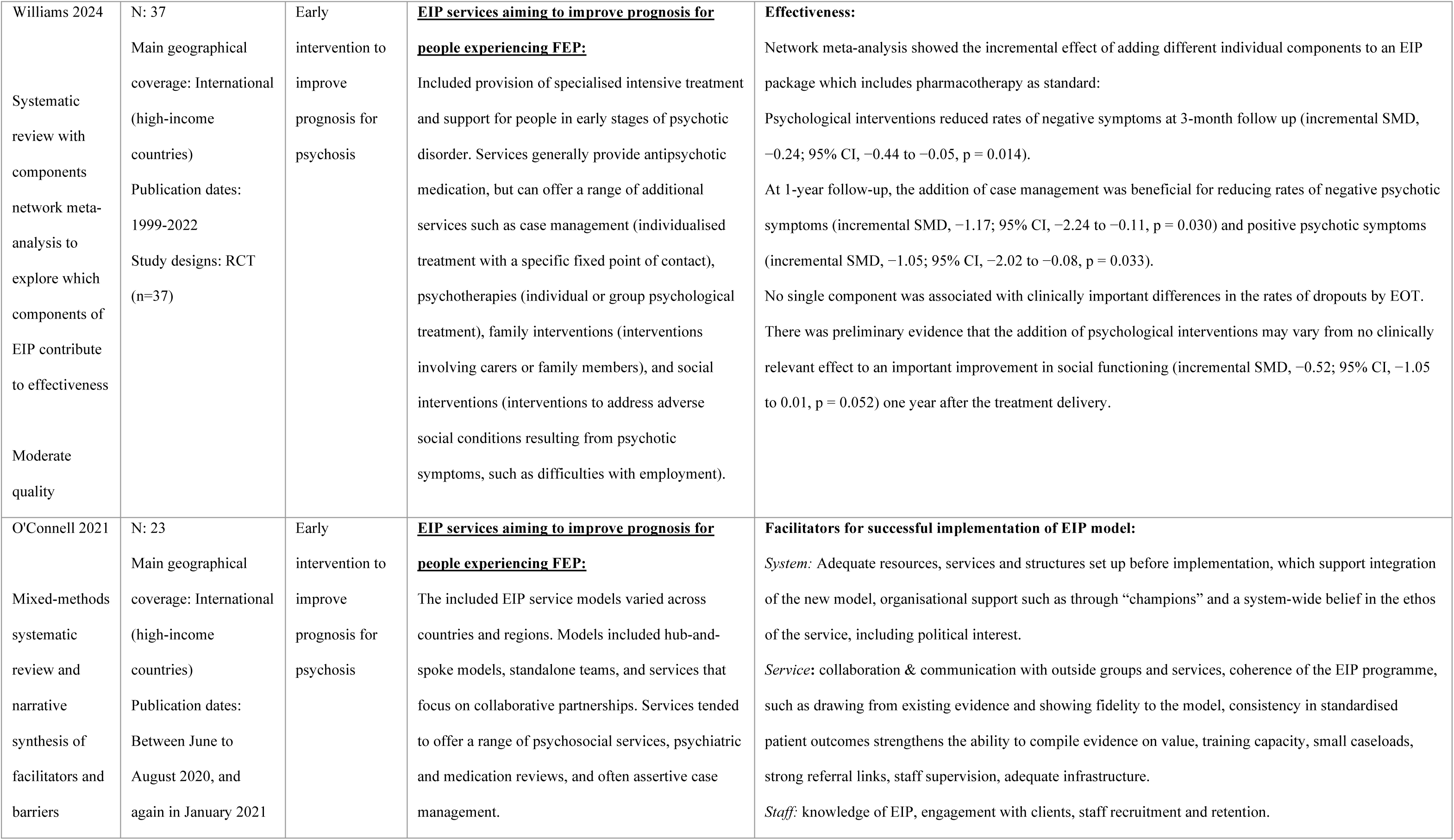
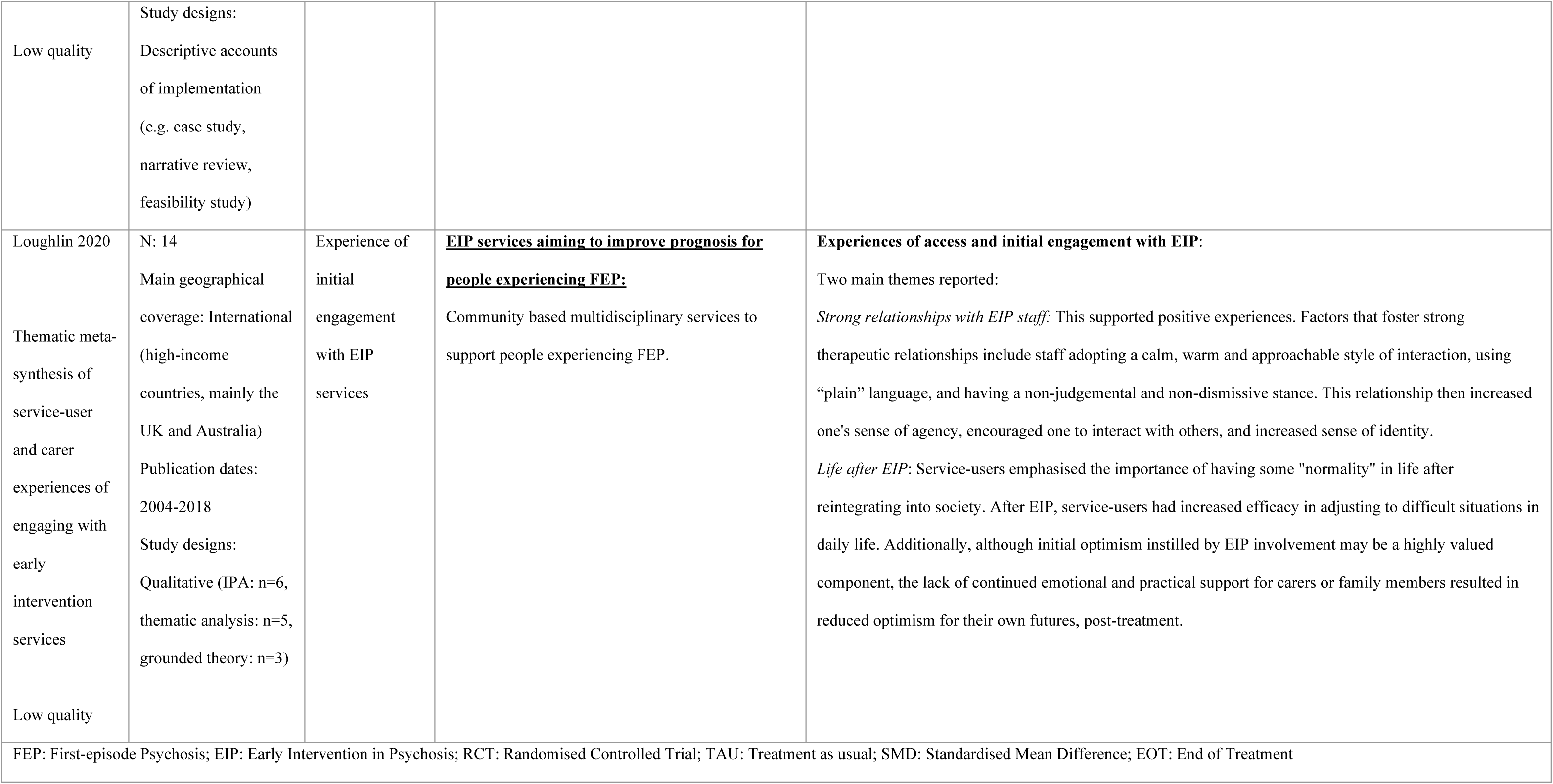
Reviews of early intervention models to improve prognosis for FEP.

##### Strategies to reduce DUP and improve pathways to care

Reviews described interventions to reduce DUP such as training healthcare professionals, particularly in primary care, to identify signs of FEP and refer patients to appropriate services (Aceituno et al., 2021), as well as multi-component public health strategies (Murden et al., 2024; Salazar de Pablo et al., 2024). These public health strategies were aimed at members of the general public experiencing early signs of FEP and their family and friends, and at professionals in healthcare and elsewhere who are likely to come into contact with people experiencing FEP. Strategies included alterations to pathways to care, awareness campaigns on the signs of FEP and available support, and educational interventions for specific groups (Murden et al., 2024).

*Effectiveness*: There was mixed evidence on the effect of models in reducing DUP (Aceituno et al., 2021; Murden et al., 2024) – although they may result in higher functioning levels at service entry (Salazar de Pablo et al., 2024), and identify people experiencing long-term symptoms (>2 years), evidence for this was mixed (Murden et al., 2024). Models with multiple targets (general public, non-health professionals and health professionals) delivered across longer periods of time may be more likely to reduce DUP (Murden et al., 2024).

Salazar de Pablo et al (Salazar de Pablo et al., 2024) also reported that all early intervention in psychosis models, whether they have reducing DUP as a primary aim or are mainly focused on improving prognosis following first contact with services, have a small overall effect on DUP (g=0.17, 95% CI: 0.06-0.28).

*Implementation*: Training healthcare staff to recognise psychosis and refer individuals earlier had good acceptability and increased skills (Aceituno et al., 2021), although lack of time and poor coordination between services were barriers to the implementation of this intervention.

Two reviews (one high- and one low-quality) synthesised evidence on barriers and facilitators to accessing early intervention services (primarily those with a main aim of improving prognosis), highlighting areas with scope to achieve further reduction in DUP. Both reviews identified as barriers to access negative perceptions of psychiatric services and medication, as well as stigma associated with seeking mental health support, or a lack of knowledge (among both patients and health professions) about key signs of psychosis (Causier et al., 2024; Tiller et al., 2023). Misalignment between available resources and patient needs also resulted in delayed access to care through short appointments and a lack of continuity of care (Causier et al., 2024). Availability of high-quality support from family and friends, collaborative and flexible services, and provision of accurate information (for example through public health campaigns) were reported as facilitators to navigating complex care systems in one low-quality review (Tiller et al., 2023).

Table 1 provides further information on reviews describing evidence on early interventions to reduce DUP and improve pathways to care.

##### Early intervention services to improve prognosis

The majority of included reviews described Early Intervention in Psychosis (EIP) service models with a main aim of improving prognosis following presentation to services. Services varied across countries and regions (O’Connell et al., 2021) but models typically included rapid access to antipsychotic medication, individual or group psychological interventions, case management, and family involvement, delivered by multidisciplinary, collaborative teams in the community. Some also described social interventions such as employment support (Farooq et al., 2024; Williams et al., 2024) and an assertive outreach style as central to service offerings (Puntis et al., 2020). EIP services reviewed in Latin American (Aceituno et al., 2021) and Low and Middle Income Countries (LMICs) (Farooq et al., 2024) described similar services, although some adaptations such as greater importance of case managers and additional provision of physical health interventions were also described (Farooq et al., 2024).

One review (Farooq et al., 2024) included a study focused on an alternative, less resource-intensive model which was considered potentially suitable to LMIC settings. This involved depot antipsychotic medication prescription alongside an assertive monitoring programme by mental health nurses to encourage continued engagement.

*Effectiveness:* A high-quality Cochrane review of EIP trials concluded with low certainty that EIP services increased likelihood of recovery, reduced admissions to psychiatric hospitals, and improved functioning. The review also concluded with moderate certainty that EIP services reduce the risk of disengagement from services at the end of treatment by half compared to treatment as usual, although general psychotic symptoms at end of treatment did not significantly differ (Puntis et al., 2020). A moderate-quality review reported that EIP models resulted in significantly larger improvements over time than usual for measures of quality of life, employment, and functioning, but that evidence regarding improvements in symptoms and remission was mixed (Salazar de Pablo et al., 2024). Moderate-quality reviews of EIP services in LMICs and Latin American countries reported that in these contexts there were fewer relapses and reduced symptomatology compared to controls in an RCT as well as over time in longitudinal studies (Aceituno et al., 2021; Farooq et al., 2024).

One moderate-quality review (Williams et al., 2024) explored which individual components of EIP services are most effective (combined with antipsychotic medication). Although psychological interventions reduced rates of negative symptoms at 3-month follow-up, at longer (12-month) follow-ups evidence of this effect was less clear. However, case management was beneficial for reducing both negative and positive symptoms, with large effect sizes.

In environments with limited resources, combining a depot antipsychotic with assertive monitoring was reported to be an effective alternative treatment model for first-episode schizophrenia (Farooq et al., 2024).

*Implementation:* Moderate-quality evidence suggested that the key components of EIP services can be adapted and provided in resource-poor settings such as LMICs (Farooq et al., 2024), and that studies in Latin America demonstrated feasibility and initial penetration (Aceituno et al., 2021), although few studies were scaled up from initial local implementation.

Facilitators of successful implementation of early intervention services to improve prognosis noted in two (moderate- and low-quality) reviews (Farooq et al., 2024; O’Connell et al., 2021) included collaboration and communication with other health services, and sufficient training capacity and supervision within teams, which in turn supported recruitment and retention of staff. Adequate funding, existing service structures, and support for the model from, for example, political leaders, were also noted in the low-quality review (O’Connell et al., 2021).

*Experiences of care:* Two moderate-high quality reviews briefly reported that satisfaction ratings were higher for patients receiving EIP services than controls (Puntis et al., 2020; Salazar de Pablo et al., 2024). Qualitative literature suggested that strong relationships with staff supported increased agency, sense of identity, and confidence to interact with others, and that early interventions supported readjustment to normal life, although the lack of continued, ongoing support following discharge reduced optimism for the future (described in one low-quality review (Loughlin et al., 2020)).

Table 2 provides further information on reviews describing early interventions to improve prognosis for FEP.

#### Early interventions for Eating Disorders

Three reviews synthesised research on early intervention approaches for eating disorders. One rapid review included evidence on models which included specialist care provision within standard mental health pathways to support identification and referral to treatment (Pehlivan et al., 2022). Another rapid review included both models aiming to reduce the duration of untreated eating disorder (DUED) and models aiming to improve prognosis once contact had been made with services (Koreshe et al., 2023), however both of these reviews were of critically low quality. One Health Technology Assessment (HTA) synthesised evidence for early interventions to improve prognosis (Hamson et al., 2023). Tables 3 and 4 describe individual review characteristics and outcomes for strategies to improve DUED and pathways to care, and strategies to improve prognosis, respectively.

**Table 3:**
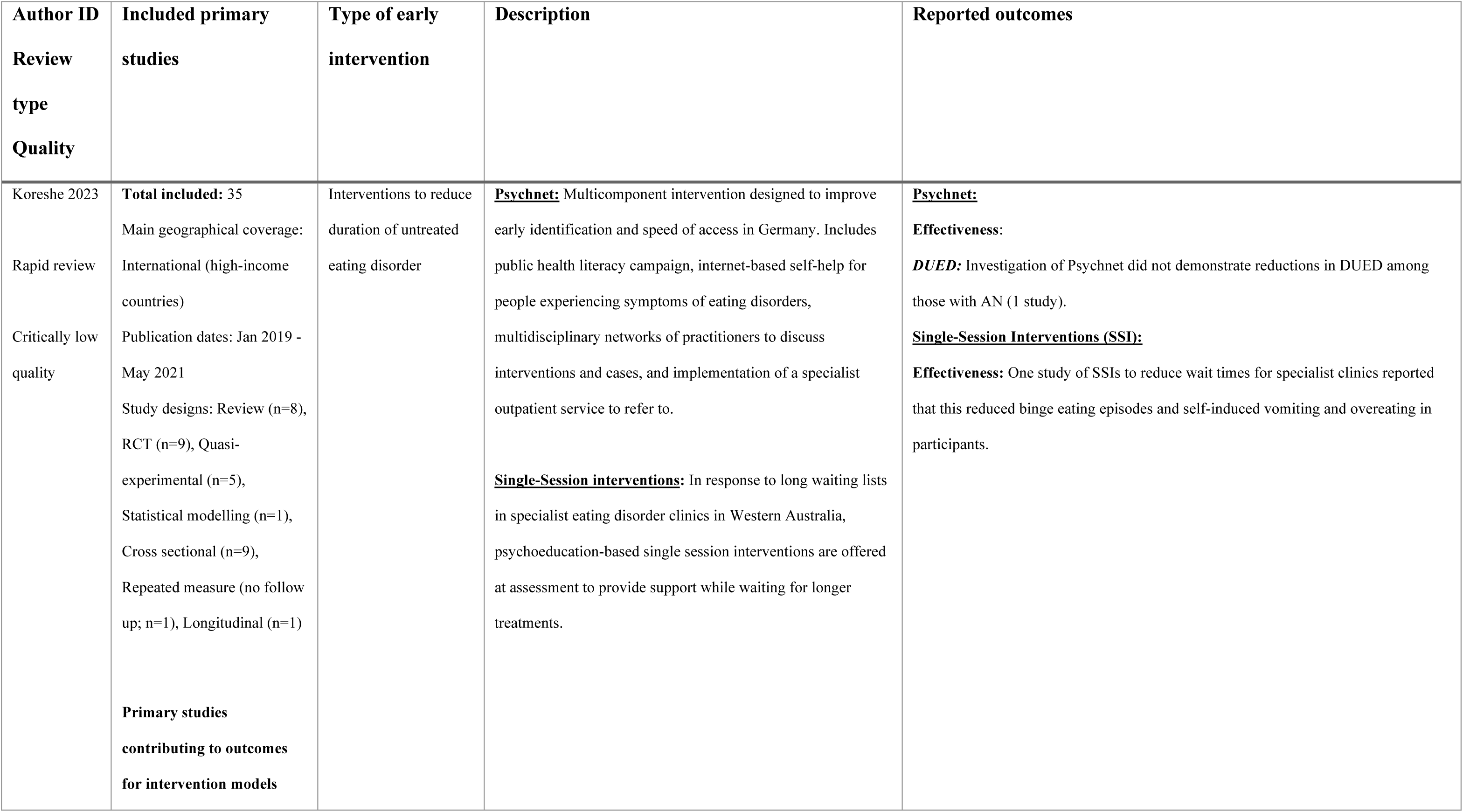
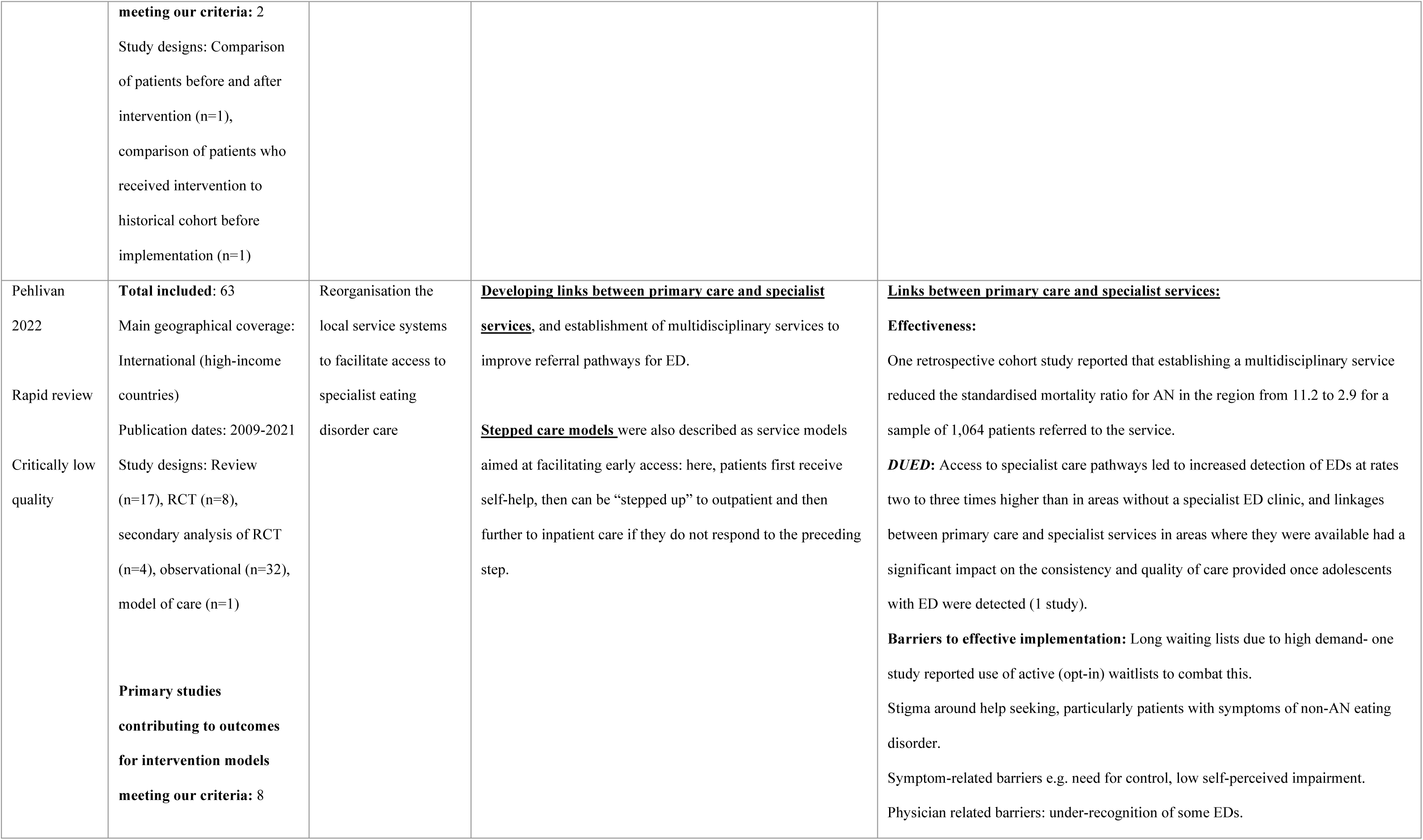
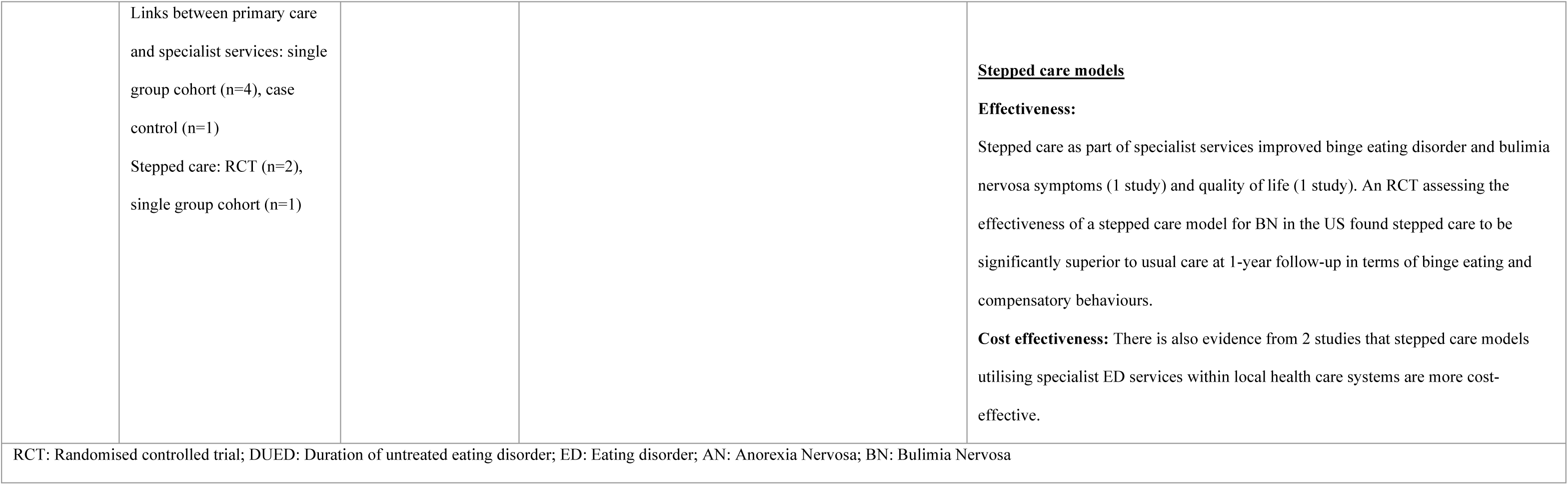
Reviews of early intervention models to reduce DUED and improve pathways to care.

**Table 4:** Reviews of early intervention models to improve prognosis for ED.

| Author<br>ID<br>Review<br>type<br>Quality | Included primary studies | Type of<br>early<br>intervention | Description | Reported outcomes |
| --- | --- | --- | --- | --- |
| Hamson<br>2023<br>HTA (no<br>meta-analysis)<br>Moderate<br>quality | **Total included:** 14<br>Main geographical coverage: International (high-income countries)<br>Publication dates: 2012-2023<br>Study designs: Cohort study (n=12), RCT (n=2)<br>**Primary studies contributing to outcomes for intervention models meeting our criteria:** 8<br>Study designs: Controlled pre-post cohort (n=6, with some overlapping samples), retrospective cohort study (n=1), single-arm pre-post cohort study (n=1) | Early<br>intervention to<br>improve<br>prognosis for<br>eating disorders | **FREED:** a UK service model in eating disorder services which aims to offer early assessment and treatment according to prespecified wait time targets alongside evidence-based treatment such as CBT, Maudsley anorexia nervosa treatment for adults, and tailoring for developmental needs and early stage of illness.<br>**Emerge-ED,** an Australian model, is modelled after FREED (n=1 study). | **Effectiveness:**<br>Findings from 7 non-randomised FREED-based studies (including 6 with a TAU comparator) and 1 single-arm non-randomised study of Emerge-ED suggest that compared to both before the intervention and TAU controls, participants who were included in early intervention program service models experienced significant reductions in eating disorder symptomology (4 studies), eating disorder cognition-related outcomes (1 study), bingeing and purging behaviour episodes (3 studies), laxative use (3 studies), excessive exercise behaviour (2 studies), and restrictive dieting behaviour (1 study). Participants provided early intervention also showed reduced psychological distress (3 studies), psychological impact due to eating disorders (3 studies), depression, anxiety, and stress (3 studies), improved function and well-being (1 study) and work and social adjustment (2 studies). Increases in mean BMI were reported up to 12-month follow-up (3 studies), or were higher than in retrospective TAU cohorts (2 studies). At longer follow-up measures, a higher proportion of FREED participants were described as weight recovered compared to TAU participants (3 studies). Overall there was a lack of comparative evidence, making interpretation of<br>estimated differences challenging.<br>**DUED:** Two FREED-based studies suggested that, when compared to a retrospective TAU cohort, those who were involved in the FREED study experienced mixed findings for duration of eating disorder onset to specialist contact (DUSC) but had lower DUED. |
| Koreshe<br>2023<br>Rapid review<br>Critically<br>low quality | **Total included:** 35<br>Main geographical coverage: International (high-income countries)<br>Publication dates: Jan 2019 - May 2021<br>Study designs: Review (n=8), RCT (n=9), Quasi-experimental (n=5), Statistical modelling (n=1), Cross sectional (n=9), Repeated measure (no follow up; n=1), Longitudinal (n=1)<br>**Primary studies contributing to outcomes for intervention models meeting our**<br>**criteria: 1**<br>Study design: Pilot RCT* | Early intervention to improve prognosis for eating disorders | **FREED:** a UK service model in eating disorder services which aims to offer early assessment and treatment according to prespecified wait time targets alongside evidence-based treatment such as CBT, Maudsley anorexia nervosa treatment for adults, and tailoring for developmental needs and early stage of illness. | **Effectiveness:**<br>Results of a FREED pilot suggested that provision of psychological treatments produced significant reductions in ED symptoms and increases in BMI.<br>**DUED:** FREED participants had a mean waiting time for treatment of 42 days compared to 62 days in the control group between referral and assessment. |
| FREED: First Episode and Rapid Early Intervention; RCT: Randomised Controlled Trial; DUED: Duration of Untreated Eating Disorder; CBT: Cognitive Behavioural Therapy<br>*This rapid review included a large number of interventions, however, detail was very limited, meaning we were only able to clearly include the FREED model, however it is possible that other interventions such as digital interventions in some instances also were integrated into more complex forms of support. |  |  |  |  |

##### Strategies to reduce DUED and improve pathways to care for people with ED

Strategies to reduce duration of untreated symptoms identified in two rapid reviews included Single session interventions (SSIs) within assessment sessions to prevent a long wait for specialist support (Koreshe et al., 2023), and multidisciplinary networks and linkages between primary care and specialist services (Koreshe et al., 2023; Pehlivan et al., 2022).

Linkages were included as part of a multi-component campaign (Psychnet) which also included internet-based self-help for people experiencing symptoms and a public health literacy campaign in one instance (Koreshe et al., 2023). Stepped care models were also discussed as a means to facilitate rapid access, where patients first receive self-help which can be provided even when symptoms are at an early stage, with subsequent “step-up” to outpatient and further to inpatient care if they do not respond to the preceding step (Pehlivan et al., 2022).

*Effectiveness:* As only two critically low-quality rapid reviews included models to reduce DUED or improve pathways to care, with few high-quality primary studies evaluating effectiveness, drawing conclusions on effectiveness is challenging. The impact of the multi-component Psychnet model was only evaluated in one small primary study, with results suggesting no reductions in DUED (Koreshe et al., 2023). However, one retrospective study demonstrated increased detection of eating disorders and improved quality of care through developing better links between primary and specialist care (Pehlivan et al., 2022). SSIs provided after assessment improved some symptoms over time (before further intervention) in one primary study (Koreshe et al., 2023), however no further information was provided. Furthermore, based on one primary study each, both specialist referral pathways to a multidisciplinary service and stepped care as part of specialist services were reported to reduce severity of AN or BN, respectively (Pehlivan et al., 2022).

*Barriers to Implementation:* One critically low-quality review (Pehlivan et al., 2022) cited long wait lists, patient-related barriers such as a need for control, lack of physician knowledge, and stigma as key barriers to accessing early interventions for eating disorders. Table 3 provides further information on reviews describing early intervention models to reduce DUED and improve pathways to care for eating disorders.

##### Early intervention services to improve prognosis for people with ED

First Episode and Rapid Early Intervention (FREED) was described in two reviews (Hamson et al., 2023; Koreshe et al., 2023). This model of early intervention originates from the UK and is delivered within some UK NHS service systems. It has a holistic and person-centred approach, providing evidence-based psychotherapy tailored to the individuals’ needs and the stage of their condition. Another early intervention service in Australia, Emerge-ED, was also described (Hamson et al., 2023) and is modelled on FREED.

*Effectiveness:* Evidence from seven non-randomised studies included in the moderate-quality HTA review (Hamson et al., 2023) and single pilot RCT included in the critically low-quality rapid review (Koreshe et al., 2023) indicated that participants who received FREED-based models experienced reduced waiting times compared to a retrospective treatment as usual (TAU) cohort as well as an improvement across a range of symptoms up to 12 months follow-up, compared to a comparison group. This included improved weight at longer follow-up points (Hamson et al., 2023; Koreshe et al., 2023). However, samples overlapped in available primary studies and evidence consisted primarily of retrospective cohort and pilot study data, making interpretation of estimated differences challenging.

Table 4 provides further information on reviews describing early interventions to improve prognosis for ED.

#### Early intervention for Bipolar Disorder

One moderate-quality review included early interventions to improve prognosis for bipolar disorder (Ratheesh et al., 2023). This review primarily included pharmacological and psychological interventions but also included one early intervention service meeting inclusion criteria, described in one primary study. The BD Specialised Mood Clinic was a service for patients discharged after their first, second, or third hospital admission for bipolar disorder, offering both pharmacological interventions and group-based psychoeducation provided by a multidisciplinary team.

*Effectiveness:* The review reported that in one included study, the risk of subsequent re-admission was found to be significantly lower in individuals treated in the specialised mood clinic (Hazard Ration (HR) = 0.60, 95% CI 0.37-0.97) compared to those in standard care (Ratheesh et al., 2023).

*Experiences:* The review reported that participants reported greater satisfaction with care in the specialised mood clinic (no further detail) (Ratheesh et al., 2023).

Further information on the review describing early intervention to improve prognosis for bipolar disorder is available in Table 5.

**Table 5:**
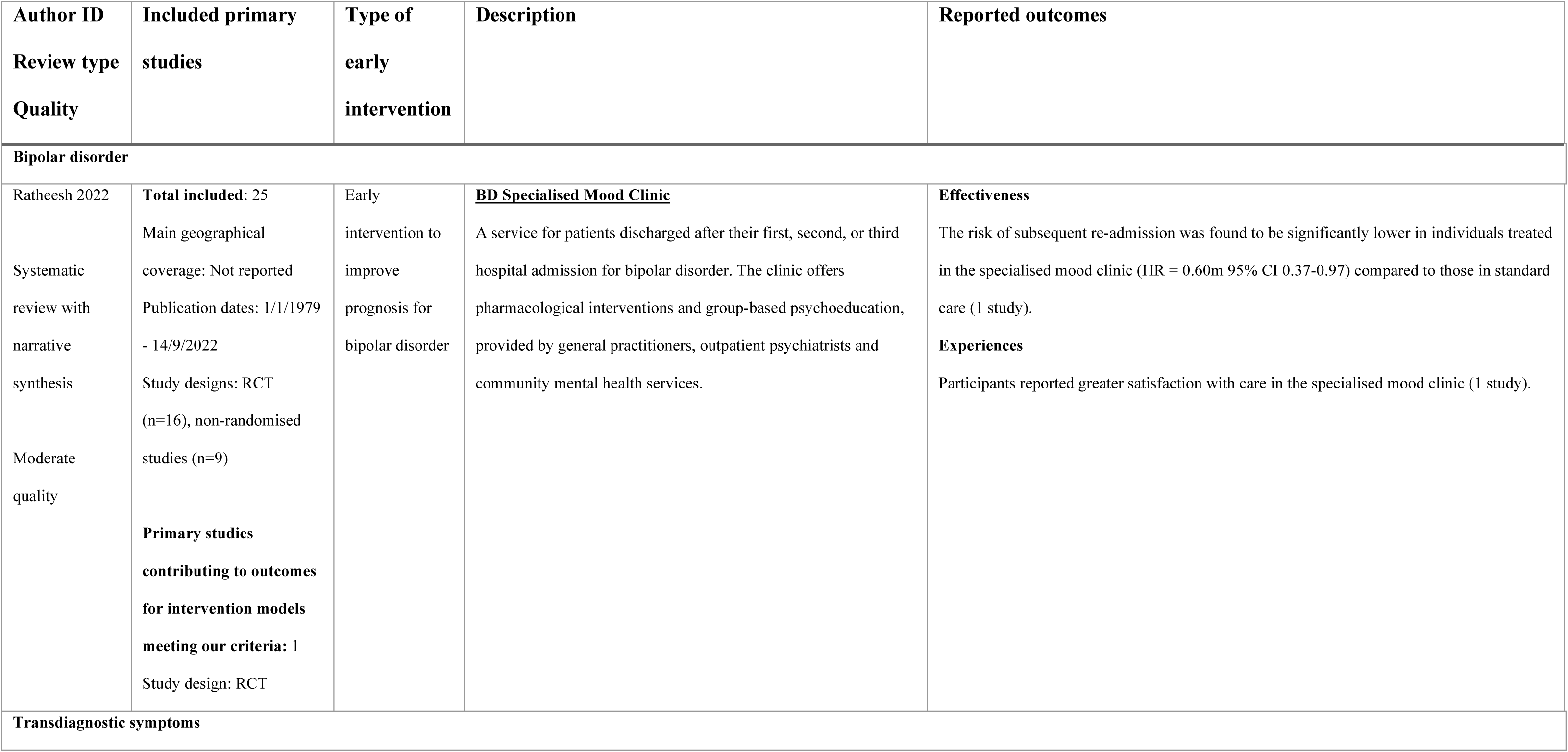
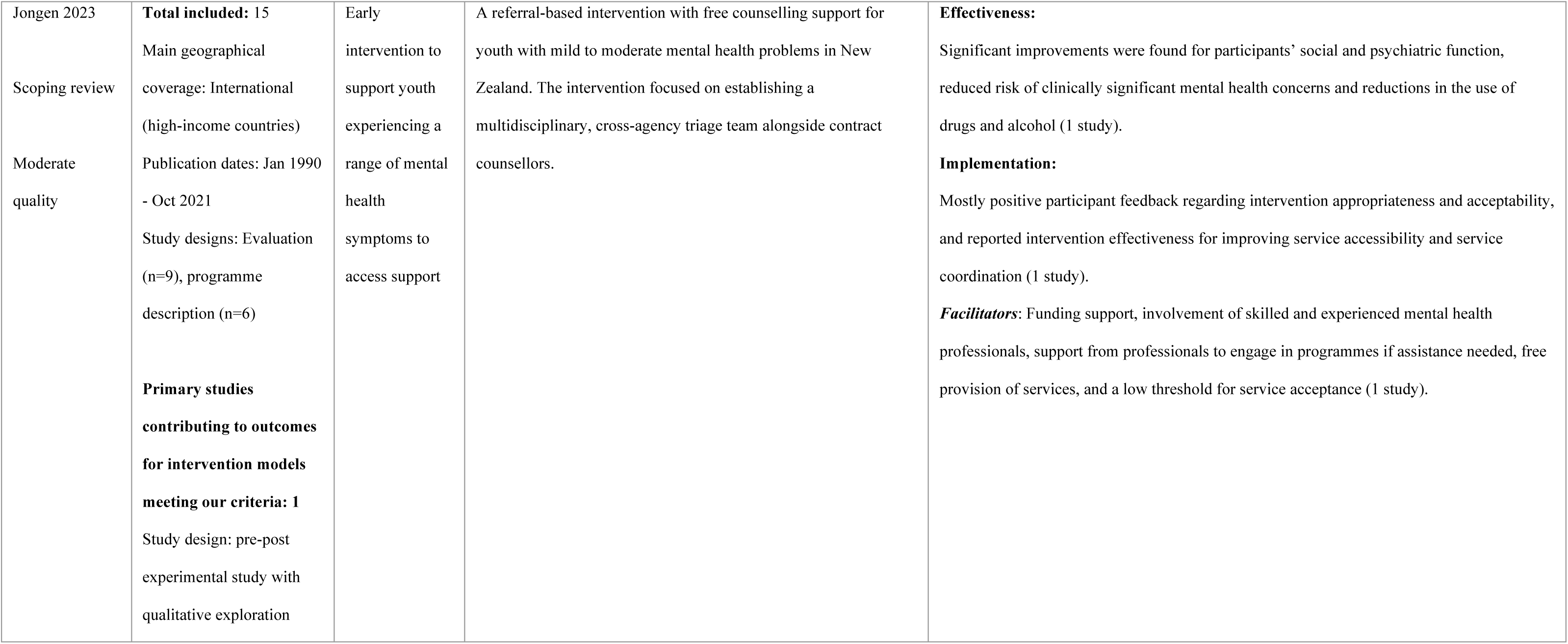
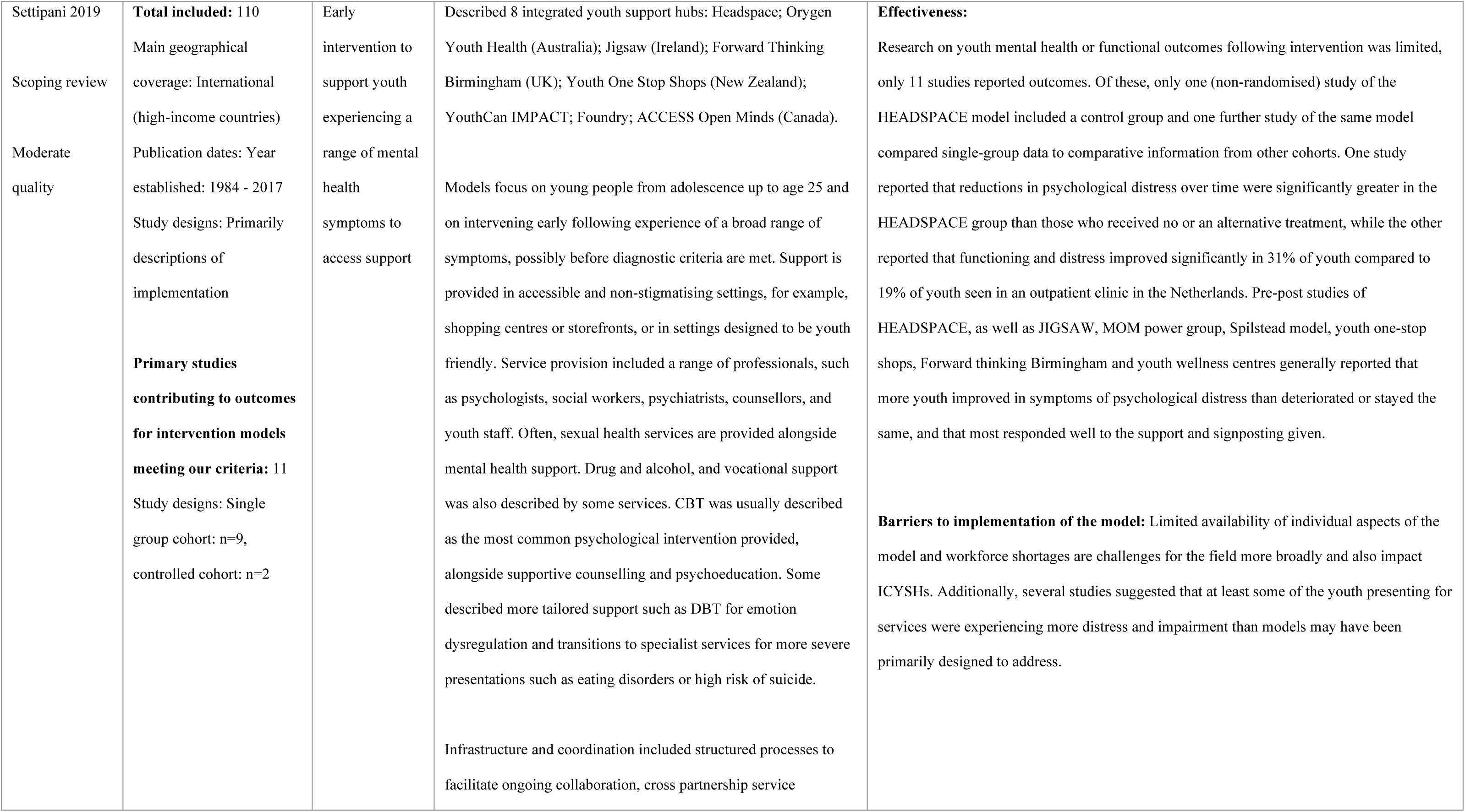
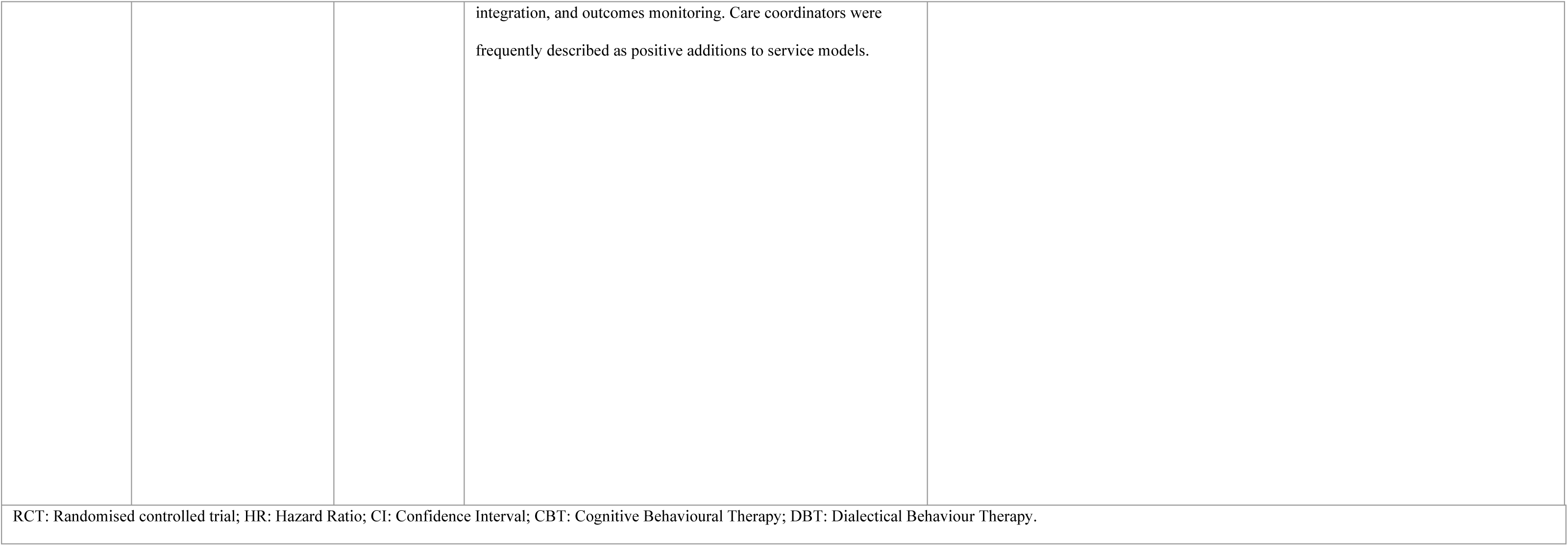
Reviews of early intervention models for bipolar disorder and transdiagnostic mental health problems.

#### Transdiagnostic early intervention models

Two reviews synthesised information relating to intervention services which did not have a specific mental health disorder focus, instead aiming to support young people with early symptoms of any mental health problem. One review synthesised information relating to integrated community-based youth hub models (ICYSHs), which provide comprehensive ‘one-stop-shop’ services for young people in community-based settings, integrating mental health services such as counselling with other community and social services such as housing support (Settipani et al., 2019). ICYSHs commonly include a multidisciplinary team and family engagement to improve service delivery. A second review of a range of interventions for indigenous youth included one additional referral-based intervention to improve access to mental health care (Jongen et al., 2023). This intervention established a multi-disciplinary triage team from youth health services, school-based services, and child and adolescent mental health services which worked to identify youth experiencing symptoms and referred them to counsellors for early intervention support (Jongen et al., 2023).

*Effectiveness:* There is limited evidence of effectiveness of transdiagnostic early intervention models, with one moderate-quality review finding that only 11 of 110 papers describing integrated community youth hubs reported effectiveness outcomes, and only two of these comparing to a control. However, the review stated that those that did report outcomes generally reported these as positive, with improvements in psychological distress and psychosocial functioning over time (Settipani et al., 2019). The referral-based intervention for youth demonstrated improvements in social and psychiatric functioning, a reduced risk of clinically significant mental health outcomes, and a decrease in the use and impact of drugs and alcohol following implementation (Jongen et al., 2023).

*Barriers and facilitators to Implementation:* Mainly positive feedback regarding intervention appropriateness and acceptability was reported by the single primary study evaluating the referral intervention, alongside improved service accessibility and coordination (Jongen et al., 2023). A low threshold for acceptance, funding support and involvement of skilled professionals were cited as facilitators.

Barriers in service implementation for ICYSHs were found to be limited service availability and a shortage of healthcare staff. Some evidence suggested that populations seen in the hubs were experiencing more distress and impairment than the model was designed to address (Settipani et al., 2019).

Further information on the reviews describing transdiagnostic early intervention models is available in Table 5.

## Discussion

This umbrella review brings together evidence for early intervention approaches across different types of mental health problems, as reported so far in systematic reviews. Overall, there was good evidence for benefits of early intervention models to improve prognosis for people experiencing symptoms of psychosis, although review-level evidence for other diagnoses and efforts to reduce duration of untreated symptoms presents a less clear picture. EIP services were reported to improve recovery and across a range of measures such as functioning, although evidence was less clear on impacts on psychotic symptom severity (Aceituno et al., 2021; Puntis et al., 2020; Salazar de Pablo et al., 2024; Williams et al., 2024). One review (Williams et al., 2024) provided novel preliminary evidence on the effectiveness of specific components of EIP, suggesting that psychological interventions and case management may be more beneficial than pharmacotherapy alone. This offers an evidence-based approach to identifying ‘essential’ components of EIP, building upon Addington et al.’s (Addington et al., 2013) work using expert consensus. The economic benefits of EIP have been highlighted across health systems, which can be attributed to reduced uptake of crisis and inpatient services and better employment outcomes (Aceituno et al., 2019; Knapp & Wong, 2020).

Evaluative evidence is very limited for early interventions meeting the criteria for complex interventions to improve prognosis in eating disorders. However, initial evaluation from observational studies of outcomes of the FREED model compared to TAU suggests that a similar holistic, multidisciplinary approach taken by EIP services may also support people experiencing early symptoms of eating disorders (Hamson et al., 2023; Koreshe et al., 2023). The importance of early management of eating disorders has been stressed in the literature, and many interventions which did not meet our criteria for complex interventions were described in these reviews (for example online interventions which may target a wider population who may not access specialist ED Services (Koreshe et al., 2023)). However, this umbrella review highlights the need for more systematically reviewed, high-quality evidence for complex early intervention services to support identification and treatment of ED symptoms.

We also found very little review evidence for early interventions to support people experiencing a range of early symptoms of common mental health problems such as anxiety and depression. Although one review highlighted a large literature base describing transdiagnostic community hub models which may be potentially promising in improving symptom severity, reducing wait times, and preventing exacerbation of symptoms (Settipani et al., 2019), there was a lack of high-quality primary evidence such as controlled studies available for such services. Given the rise in prevalence of anxiety and depression among young people in the last few decades globally (Cao et al., 2024; Javaid et al., 2023; Shorey et al., 2022), there is also a need for the current evidence base, which consists of primarily single-group evaluations, to be supplemented with controlled comparative studies. Finally, we found almost no evidence for early intervention approaches for other mental health conditions, such as bipolar disorder, where we found only one model of complex early intervention described within one review (Ratheesh et al., 2023).

Although early interventions to improve prognosis such as EIP can also include targeted efforts to provide treatment quickly, resulting in positive though small effects on DUP (Salazar de Pablo et al., 2024), review evidence for approaches specifically designed to reduce duration of untreated symptoms is mixed, although particularly limited for non-psychosis presentations (Aceituno et al., 2019; Koreshe et al., 2023; Murden et al., 2024; Salazar de Pablo et al., 2024). This may be because there are substantial methodological challenges in carrying out studies in this area, for example, randomisation for public health campaigns or stigma reduction is generally not feasible. Despite this, some reviewed evidence has demonstrated varying levels of success in including a range of intervention components to support early identification, including public education, stigma reduction, and improvement in connections between services. Further investigation is needed to explore which are most effective and how best to achieve sustained implementation. Relatedly, knowledgeable healthcare professionals and ensuring availability of services to support rapid referrals were reported as facilitators to successful early intervention across diagnoses (Aceituno et al., 2019; O’Connell et al., 2021; Pehlivan et al., 2022).

In line with a large evidence base that stigma significantly affects access to a broad range of mental health support (Clement et al., 2015; Schomerus & Angermeyer, 2008), reviews reported that negative perceptions of services alongside societal and personal stigma impacted access to early interventions, particularly for psychosis and eating disorders. Lack of resources, such as sufficient specialist services to meet demand or adequately trained healthcare staff, also impeded access for those experiencing symptoms of eating disorders and common mental health problems (Pehlivan et al., 2022; Settipani et al., 2019). This is in line with previous calls for additional funding in this area (Every-Palmer et al., 2024; O’Connor et al., 2023), particularly regarding support hubs for young people (McGorry et al., 2022). Findings also suggested that lack of knowledge regarding the nature of symptoms and ways to seek help could prevent timely access to care. Support from family and friends was reported to combat this by facilitating navigation of complex care systems in one review of EIP (Tiller et al., 2023), supporting previous qualitative evidence on the role of family in identifying symptoms and subsequently, available support (Oluwoye et al., 2020).

Few reviews reported experiences of care although the available evidence suggested that patients are more satisfied with early intervention efforts than traditional treatment services (Puntis et al., 2020; Ratheesh et al., 2023), and that early intervention in psychosis can contribute to improved agency and re-integration within society after the end of treatment (Loughlin et al., 2020). The reported experiences of service users in their lives after EIP (for example the importance of social reintegration, and increased ability to handle difficult situations) have been deemed among the most important outcomes of general treatments for psychosis by service users (Byrne & Rosen, 2014). This stresses the effectiveness of early intervention in psychosis from a service-user perspective. Despite this, it would be of benefit to further explore specifically how early intervention models may improve experiences of mental health support.

### Strengths and limitations

This systematic umbrella review provides a broad overview of the state of the evidence for early intervention approaches across a number of symptom presentations, including the impact of these services on effectiveness, experiences, and implementation, highlighting current gaps in the evidence base. These evidence gaps are significant in most instances, primarily due to a lack of primary research comparing early intervention models to controls for most diagnoses, limiting conclusions that can be drawn. We did not find systematically reviewed evidence for early intervention strategies for behaviours and difficulties resulting in a personality disorder diagnosis. It has been suggested that young people presenting to care with these difficulties may not be identified, or that many clinicians believe that a diagnosis of a personality disorder necessitates specialist psychotherapy programmes which cannot be accessed rapidly (Chanen et al., 2022), thus may inhibit the development of early interventions in this field.

Umbrella reviews by definition also seek to answer broader research questions through synthesis of syntheses (Gianfredi et al., 2022), necessitating a lack of detailed focus on individual primary research, such as specific intervention protocols which may vary between individual primary studies. Umbrella reviews also entail a time-lag in evidence synthesis (Gianfredi et al., 2022) which may mean that some recent high-quality research in this area has not been summarised here as it has not been reported in reviews.

### Implications for research, policy and practice

Results suggest that early intervention models are effective in improving prognosis for people experiencing symptoms of psychosis. EIP models which are individualised, multidisciplinary and provide rapid access to evidence-based care have a substantial evidence base and therefore, effective implementation of these approaches should be considered a priority.

Although evidence included in systematic reviews was markedly lacking in controlled effectiveness studies for most other early intervention approaches, alongside limited longer-term exploration of impact, both are important to understand the true economic and societal impacts of early interventions and should be a priority for both primary research and review syntheses in the future to support provision of care across mental health conditions and prevent further exacerbation of symptoms. While one recent RCT for an early intervention model for bipolar disorder (Ratheesh et al., 2023) and some early controlled evaluations for the FREED model for eating disorders (Hamson et al., 2023) were included in reviews, our results nevertheless highlight the need for further exploration of the effectiveness of early intervention approaches for eating disorders, bipolar disorder, depression, anxiety disorders, and behaviours and difficulties resulting in a personality disorder diagnosis. Furthermore, there is little available evidence at present for effective approaches to reduce duration of untreated illness, although integration of specialist support with clear pathways for referral has been highlighted as potentially helpful. This is a clear target for additional primary research which could further consider the barriers faced by those experiencing early symptoms in accessing care. Such work should also seek to have greater involvement of researchers with lived experience, which was limited in currently available syntheses.

## Conclusions

Overall, evidence suggests that early intervention approaches can improve outcomes for people experiencing early symptoms of psychosis. While evidence for early intervention in other diagnoses is limited, initial studies point towards benefits in improving access and symptom severity, although further high-quality comparative studies are required. Efforts to improve identification and access to support may offer some benefit, however, further exploration is needed to determine how best to reduce the duration of untreated symptoms. Integrating these efforts with other available early interventions options could be most effective. Models which combat limited resources through linkages and collaboration alongside staff training and dissemination of information for service users and families could be a key facilitator of successful implementation.

## Lived experience commentary

### Written by two members of our working group with lived experience: Lizzie and Eva

We are a young person with lived experience and a carer of young people with mental health conditions and long-term engagement with CAMHS.

We welcome this much-needed review into current approaches for early intervention (EI) services for children and young people (CYP). We are disappointed by the lack of published evidence in this area, and the low quality and lack of scientific rigour in the studies examined.

Our initial question from this research is how can we intervene early if we are not treating the first stages of an emerging mental illness?

The studies reviewed are for *more serious mental illnesses* such as psychosis and eating disorders led by psychiatric, diagnostic, medical models, whereas we feel CYP could benefit from more needs-led, not diagnosis-led approaches.

There is a lack of research into early intervention for the more common problems such as depression and anxiety, which in our experience with CYP mental health care, can present as early warning signs and can lead to serious educational and vocational problems and also be precursors to more severe mental ill-health. It is clearly difficult to draw conclusions from the evidence examined and more robust work with a broader range of presentations and symptoms is desperately needed.

Every young person deserves access to an early intervention approach for all signs of mental distress, including anxiety and depression. The care needs to be flexible in approach, location, time frame, and personalised to the needs of the young person, with extra consideration given for easy-to-ignore populations due to language, culture, economic circumstances and those in rural locations who can’t just ‘drop in’ to a city-centre hub as these are heavily skewed towards urban areas.

Mental health difficulties don’t end when a person leaves the therapy room and neither should mental health support. Home or school visits, outreach, practical support, and a ‘triangle of care model’ equally considering CYP, carers and family, and the professionals views, can help mental health support be independently accessible, create a safe and stigma-free environment and empower the young person to create sustainable improvements in their mental health.

The Early Support Hubs are a new model of care aiming to adopt the more needs-led, collaborative and accessible approaches CYP need for mild-moderate mental health difficulties such as anxiety and depression, to address the aforementioned gaps in current EI services. We hope this will fulfil the ‘early’ part of the ‘early intervention’ promise, providing the effective, proactive and accessible support young people so desperately need for emerging signs of mental distress.

## Data Availability

All data produced in the present study are available upon reasonable request to the authors

## Funding

This work was commissioned and funded by the National Institute for Health and Care Research (NIHR) Policy Research Programme and conducted by the NIHR Policy Research Unit in Mental Health (MHPRU). The funder had no role in the study design, analysis, write up of the manuscript, or the decision to submit for publication.

## Availability of data and materials

The data used and/or analysed in the current study are available from the corresponding author upon reasonable request.

## Ethics Declarations

Ethics approval and consent to participate Not applicable – Umbrella Review.

## Notes

### Competing Interest Statement

The authors have declared no competing interest.

