## Appendices for "Early interventions for first onset of symptoms of mental health conditions: an umbrella review of systematic reviews"

#### Appendix 1: PRISMA Checklist

| Section and Topic | Item # | Checklist item | Location where item is reported |
|---|---|---|---|
| **TITLE** | | | |
| Title | 1 | Identify the report as a systematic review. | Title |
| **ABSTRACT** | | | |
| Abstract | 2 | See the PRISMA 2020 for Abstracts checklist. | Abstract |
| **INTRODUCTION** | | | |
| Rationale | 3 | Describe the rationale for the review in the context of existing knowledge. | Introduction |
| Objectives | 4 | Provide an explicit statement of the objective(s) or question(s) the review addresses. | Introduction |
| **METHODS** | | | |
| Eligibility criteria | 5 | Specify the inclusion and exclusion criteria for the review and how studies were grouped for the syntheses. | Methods: Eligibility criteria; evidence synthesis |
| Information sources | 6 | Specify all databases, registers, websites, organisations, reference lists and other sources searched or consulted to identify studies. Specify the date when each source was last searched or consulted. | Methods: Search strategy |
| Search strategy | 7 | Present the full search strategies for all databases, registers and websites, including any filters and limits used. | Appendix II |
| Selection process | 8 | Specify the methods used to decide whether a study met the inclusion criteria of the review, including how many reviewers screened each record and each report retrieved, whether they worked independently, and if applicable, details of automation tools used in the process. | Methods: Screening |
| Data collection process | 9 | Specify the methods used to collect data from reports, including how many reviewers collected data from each report, whether they worked independently, any processes for obtaining or confirming data from study investigators, and if applicable, details of automation tools used in the process. | Methods: Data extraction |
| Data items | 10a | List and define all outcomes for which data were sought. Specify whether all results that were compatible with each outcome domain in each study were sought (e.g. for all measures, time points, analyses), and if not, the methods used to decide which results to collect. | Methods: Eligibility criteria and Methods: |

| Section and Topic | Item # | Checklist item | Location where item is reported |
|---|---|---|---|
| | | | data extraction |
| | 10b | List and define all other variables for which data were sought (e.g. participant and intervention characteristics, funding sources). Describe any assumptions made about any missing or unclear information. | Methods: Data extraction |
| Study risk of bias assessment | 11 | Specify the methods used to assess risk of bias in the included studies, including details of the tool(s) used, how many reviewers assessed each study and whether they worked independently, and if applicable, details of automation tools used in the process. | Methods: Quality appraisal |
| Effect measures | 12 | Specify for each outcome the effect measure(s) (e.g. risk ratio, mean difference) used in the synthesis or presentation of results. | NA |
| Synthesis methods | 13a | Describe the processes used to decide which studies were eligible for each synthesis (e.g. tabulating the study intervention characteristics and comparing against the planned groups for each synthesis (item #5)). | Methods: Evidence synthesis |
| | 13b | Describe any methods required to prepare the data for presentation or synthesis, such as handling of missing summary statistics, or data conversions. | NA |
| | 13c | Describe any methods used to tabulate or visually display results of individual studies and syntheses. | Methods: Evidence synthesis |
| | 13d | Describe any methods used to synthesize results and provide a rationale for the choice(s). If meta-analysis was performed, describe the model(s), method(s) to identify the presence and extent of statistical heterogeneity, and software package(s) used. | Methods: Evidence synthesis |
| | 13e | Describe any methods used to explore possible causes of heterogeneity among study results (e.g. subgroup analysis, meta-regression). | Methods: Evidence synthesis |
| | 13f | Describe any sensitivity analyses conducted to assess robustness of the synthesized results. | NA |
| Reporting bias assessment | 14 | Describe any methods used to assess risk of bias due to missing results in a synthesis (arising from reporting biases). | NA |
| Certainty assessment | 15 | Describe any methods used to assess certainty (or confidence) in the body of evidence for an outcome. | NA |
| **RESULTS** | | | |
| Study selection | 16a | Describe the results of the search and selection process, from the number of records identified in the search to the number of studies included in the review, ideally using a flow diagram. | Results, Figure 1 |
| | 16b | Cite studies that might appear to meet the inclusion criteria, but which were excluded, and explain why they were excluded. | Appendix 4 |

| Section and Topic | Item # | Checklist item | Location where item is reported |
|---|---|---|---|
| Study characteristics | 17 | Cite each included study and present its characteristics. | Table 1 |
| Risk of bias in studies | 18 | Present assessments of risk of bias for each included study. | Table 1 |
| Results of individual studies | 19 | For all outcomes, present, for each study: (a) summary statistics for each group (where appropriate) and (b) an effect estimate and its precision (e.g. confidence/credible interval), ideally using structured tables or plots. | NA |
| Results of syntheses | 20a | For each synthesis, briefly summarise the characteristics and risk of bias among contributing studies. | Quality of included studies, table 1, Data synthesis |
| | 20b | Present results of all statistical syntheses conducted. If meta-analysis was done, present for each the summary estimate and its precision (e.g. confidence/credible interval) and measures of statistical heterogeneity. If comparing groups, describe the direction of the effect. | Results, tables 2-4 |
| | 20c | Present results of all investigations of possible causes of heterogeneity among study results. | NA |
| | 20d | Present results of all sensitivity analyses conducted to assess the robustness of the synthesized results. | NA |
| Reporting biases | 21 | Present assessments of risk of bias due to missing results (arising from reporting biases) for each synthesis assessed. | NA |
| Certainty of evidence | 22 | Present assessments of certainty (or confidence) in the body of evidence for each outcome assessed. | NA |
| **DISCUSSION** | | | |
| Discussion | 23a | Provide a general interpretation of the results in the context of other evidence. | Discussion |
| | 23b | Discuss any limitations of the evidence included in the review. | Discussion: Strengths and limitations |
| | 23c | Discuss any limitations of the review processes used. | Discussion: Strengths and limitations |
| | 23d | Discuss implications of the results for practice, policy, and future research. | Discussion: implications for research, |

| Section and Topic | Item # | Checklist item | Location where item is reported |
|---|---|---|---|
| | | | policy and practice |
| **OTHER INFORMATION** | | | |
| Registration and protocol | 24a | Provide registration information for the review, including register name and registration number, or state that the review was not registered. | Methods |
| | 24b | Indicate where the review protocol can be accessed, or state that a protocol was not prepared. | Methods |
| | 24c | Describe and explain any amendments to information provided at registration or in the protocol. | Methods |
| Support | 25 | Describe sources of financial or non-financial support for the review, and the role of the funders or sponsors in the review. | Funding |
| Competing interests | 26 | Declare any competing interests of review authors. | NA |
| Availability of data, code and other materials | 27 | Report which of the following are publicly available and where they can be found: template data collection forms; data extracted from included studies; data used for all analyses; analytic code; any other materials used in the review. | Availability of data and materials |

*From:* Page MJ, McKenzie JE, Bossuyt PM, Boutron I, Hoffmann TC, Mulrow CD, et al. The PRISMA 2020 statement: an updated guideline for reporting systematic reviews. BMJ 2021;372:n71. doi: 10.1136/bmj.n71

### Appendix 2: Search strategy

**Embase** <1974 to 2024 April 10>

1 exp mental health/ 254824
2 exp anxiety disorder/ 333874
3 exp eating disorder/ 67149
4 exp emotional disorder/ 23629
5 exp dissociative disorder/ 11424
6 exp mood disorder/ 700765
7 exp personality disorder/ 70149
8 exp psychosis/ 333340
9 exp schizophrenia spectrum disorder/ 216884
10 exp psychotrauma/ 12072
11 (mood disorder* or affective disorder* or depression or depressive or dysthymi* or anxiety disorder* or agoraphobia or obsess* or compulsi* or panic or phobi* or ptsd or posttrauma* or post trauma* or affective symptoms or personality disorder* or psychos#s or psychotic or schizophrenia or eating disorder* or anorexia or bulimia).ti,kf. 611904
12 ((mental* or psyc*) adj2 (health or well* or condition* or distress or disorder or ill* or problem* or disease* or issue*)).ti,ab,kf. 551938
13 ("at risk mental state" or "at-risk mental state" or "at-risk of psychosis" or prodrom*).ti,ab,kf. 21028
14 1 or 2 or 3 or 4 or 5 or 6 or 7 or 8 or 9 or 10 or 11 or 12 or 13 1690529
15 ((pathway* adj3 (care or mental or psyc* or service* or model or referral* or help or contact*)) or ((helpseek* or help-seek* or help seek*) adj3 (contact* or experienc* or step* or delay or duration)) or ((Referral* or treatment or "mental health service*") adj2 (pattern* or delay)) or (contact adj3 (service* or professional*)) or "Journey of care" or "point* of entry" or (entry adj2 (care or service* or treatment)) or "early intervention" or ((integrat* or multidisciplinary or multi-disciplinary or coordinat* or co-ordinat* or complex) adj2 (care or treatment or therapy or team* or service or approach or intervention)) or "early support" or "improv* access" or ((fast or faster or early) adj2 (access or referral* or signpost* or support or "mental health" or treatment)) or "support hub*").ti,kw. 81491
16 ((pathway* adj3 (care or mental or psyc* or service* or model or referral* or help or contact*)) or ((helpseek* or help-seek* or help seek*) adj3 (contact* or experienc* or step* or delay or duration)) or ((Referral* or treatment or "mental health service*") adj2 (pattern* or delay)) or (contact adj3 (service* or professional*)) or "Journey of care" or "point* of entry" or (entry adj2 (care or service* or treatment)) or "early intervention" or ((integrat* or multidisciplinary or multi-disciplinary or coordinat* or co-ordinat* or complex) adj2 (care or treatment or therapy or team* or service* or approach or intervention)) or "early support" or "improv* access" or ((fast or faster or early) adj2 (access or referral* or signpost* or support or "mental health" or treatment)) or "support hub*").ab. /freq=2 70860
17 early intervention/ or preventative mental health services/ 35123
18 (indicated adj (prevention? or intervention? or support or help or care or approach or approaches or service? or program*)).ti,kf. 169
19 ("early intervention" or "early detection" or "early support").ti. 22963
20 15 or 16 or 17 or 18 or 19 178319
21 forensic psychiatry/ or forensic psychology/ or hospital patient/ or hospitalized adolescent/ or hospitalized child/ or hospitalized infant/ 259590
22 (forensic or inpatient* or compulsor* detention or compulsor* detained or prison* or jail*).ti,kf. 113038
23 21 or 22 332921

24 20 not 23 172576
25 14 and 24 23099
26 (systematic or structured or evidence).ti. and ((review or overview or look or examination or update* or summary).ti. or review.pt.) 346925
27 meta-analysis.pt. or (meta-analys* or meta analys* or metaanalys* or meta synth* or meta-synth* or metasynth*).ti,ab,kf,hw. 479740
28 ((systematic or meta) adj2 (analys* or review)).ti,kf. or ((systematic* or quantitativ* or methodologic* or qualitativ*) adj5 (review* or overview*)).ti,ab,kf,sh. or ((quantitativ$ or qualitativ$) adj5 synthesis$).ti,ab,kf,hw. 652935
29 (evidence adj3 review*).ti,ab,kf. or "scoping review?".ti,kf. or (realist adj3 review*).ti,ab,kf. or (implementation adj3 review*).ti,ab,kf. 105520
30 26 or 27 or 28 or 29 879754
31 25 and 30 1556
32 limit 31 to ("remove preprint records" and yr="2019 -Current") 761

Ovid **MEDLINE**(R) ALL <1946 to April 10, 2024>
1 exp mental health/ 66620
2 exp anxiety disorders/ 93735
3 exp dissociative disorders/ 4847
4 exp "feeding and eating disorders"/ 37266
5 exp mood disorders/ 173151
6 exp personality disorders/ 45868
7 exp "Schizophrenia Spectrum and Other Psychotic Disorders"/ 165647
8 exp "Trauma and Stressor Related Disorders"/ 51610
9 (mood disorder* or affective disorder* or depression or depressive or dysthymi* or anxiety disorder* or agoraphobia or obsess* or compulsi* or panic or phobi* or ptsd or posttrauma* or post trauma* or affective symptoms or personality disorder* or psychos#s or psychotic or schizophrenia or eating disorder* or anorexia or bulimia).ti,kf. 468800
10 ((mental* or psyc*) adj2 (health or well* or condition* or distress or disorder or ill or problem* or disease* or issue*)).ti,ab,kf. 402445
11 ("at risk mental state" or "at-risk mental state" or "at-risk of psychosis" or prodrom*).ti,ab,kf. 12723
12 1 or 2 or 3 or 4 or 5 or 6 or 7 or 8 or 9 or 10 or 11 1009517
13 ((pathway* adj3 (care or mental or psyc* or service* or model or referral* or help or contact*)) or ((helpseek* or help-seek* or help seek*) adj3 (contact* or experienc* or step* or delay or duration)) or ((Referral* or treatment or "mental health service*") adj2 (pattern* or delay)) or (contact adj3 (service* or professional*)) or "Journey of care" or "point* of entry" or (entry adj2 (care or service* or treatment)) or "early intervention" or ((integrat* or multidisciplinary or multi- disciplinary or coordinat* or co-ordinat* or complex) adj2 (care or treatment or therapy or team* or service or approach or intervention)) or "early support" or "improv* access" or ((fast or faster or early) adj2 (access or referral* or signpost* or support or "mental health" or treatment)) or "support hub*").ti,kw. 57653
14 ((pathway* adj3 (care or mental or psyc* or service* or model or referral* or help or contact*)) or ((helpseek* or help-seek* or help seek*) adj3 (contact* or experienc* or step* or delay or duration)) or ((Referral* or treatment or "mental health service*") adj2 (pattern* or delay)) or (contact adj3 (service* or professional*)) or "Journey of care" or "point* of entry" or (entry adj2 (care or service* or treatment)) or "early intervention" or ((integrat* or multidisciplinary or multi- disciplinary or coordinat* or co-ordinat* or complex) adj2 (care or treatment or therapy or team* or service* or approach or intervention)) or "early support" or

"improv* access" or ((fast or faster or early) adj2 (access or referral* or signpost* or support or "mental health" or treatment)) or "support hub*").ab. /freq=2 42178
15 early intervention/ or preventative mental health services/ 0
16 (indicated adj (prevention? or intervention? or support or help or care or approach or approaches or service? or program*)).ti,kf. 129
17 ("early intervention" or "early detection" or "early support").ti. 16740
18 13 or 14 or 15 or 16 or 17 101027
19 forensic psychiatry/ or forensic psychology/ or hospital patient/ or hospitalized adolescent/ or hospitalized child/ or hospitalized infant/ 17415
20 (forensic or inpatient* or compulsor* detention or compulsor* detained or prison* or jail*).ti,kf. 79334
21 19 or 20 93364
22 18 not 21 100312
23 (systematic or structured or evidence).ti. and ((review or overview or look or examination or update* or summary).ti. or review.pt.) 302580
24 meta-analysis.pt. or (meta-analys* or meta analys* or metaanalys* or meta synth* or meta-synth* or metasynth*).ti,ab,kf,hw. 334771
25 ((systematic or meta) adj2 (analys* or review)).ti,kf. or ((systematic* or quantitativ* or methodologic* or qualitative) adj5 (review* or overview*)).ti,ab,kf,sh. or ((quantitativ$ or qualitativ$) adj5 synthesis$).ti,ab,kf,hw. 474436
26 (evidence adj3 review*).ti,ab,kf. or "scoping review?".ti,kf. or (realist adj3 review*).ti,ab,kf. or (implementation adj3 review*).ti,ab,kf. 95788
27 23 or 24 or 25 or 26 635934
28 22 and 27 4377
29 limit 28 to (yr="2019 -Current" and "remove preprint records") 2334

APA **PsycInfo** <1806 to April Week 1 2024>
1 exp mental health/ 97475
2 exp Anxiety Disorders/ 44238
3 exp Affective Disorders/ 182074
4 exp Bipolar Disorder/ 35489
5 exp Borderline States/ 4156
6 exp Eating Disorders/ 37046
7 exp Obsessive Compulsive Disorder/ 19103
8 exp Personality Disorders/ 32847
9 exp Psychosis/ 131534
10 exp Serious Mental Illness/ 6826
11 exp "stress and trauma related disorders"/ 44239
12 exp Dissociative Disorders/ 5904
13 (mood disorder* or affective disorder* or depression or depressive or dysthymi* or anxiety disorder* or agoraphobia or obsess* or compulsi* or panic or phobi* or ptsd or posttrauma* or post trauma* or affective symptoms or personality disorder* or psychos#s or psychotic or schizophrenia or eating disorder* or anorexia or bulimia).ti,id. 454231
14 ((mental* or psyc*) adj2 (health or well* or condition* or distress or disorder or ill* or problem* or disease* or issue*)).ti,ab,id. 577136
15 ("at risk mental state" or "at-risk mental state" or "at-risk of psychosis" or prodrom*).ti,ab,id. 6618
16 or/1-15 1007093
17 ((pathway* adj3 (care or mental or psyc* or service* or model or referral* or help or contact*)) or ((helpseek* or help-seek* or help seek*) adj3 (contact* or experienc* or step* or delay or duration)) or ((Referral* or treatment or "mental health service*") adj2 (pattern* or

delay)) or (contact adj3 (service* or professional*)) or "Journey of care" or "point* of entry" or (entry adj2 (care or service* or treatment)) or "early intervention" or ((integrat* or multidisciplinary or multi- disciplinary or coordinat* or co-ordinat* or complex) adj2 (care or treatment or therapy or team* or service or approach or intervention)) or "early support" or "improv* access" or ((fast or faster or early) adj2 (access or referral* or signpost* or support or "mental health" or treatment)) or "support hub*").ti,id. 25721
18 ((pathway* adj3 (care or mental or psyc* or service* or model or referral* or help or contact*)) or ((helpseek* or help-seek* or help seek*) adj3 (contact* or experienc* or step* or delay or duration)) or ((Referral* or treatment or "mental health service*") adj2 (pattern* or delay)) or (contact adj3 (service* or professional*)) or "Journey of care" or "point* of entry" or (entry adj2 (care or service* or treatment)) or "early intervention" or ((integrat* or multidisciplinary or multi-disciplinary or coordinat* or co-ordinat* or complex) adj2 (care or treatment or therapy or team* or service* or approach or intervention)) or "early support" or "improv* access" or ((fast or faster or early) adj2 (access or referral* or signpost* or support or "mental health" or treatment)) or "support hub*").ab. /freq=2 14265
19 early intervention/ or preventative mental health services/ 13030
20 (indicated adj (prevention? or intervention? or support or help or care or approach or approaches or service? or program*)).ti,id. 131
21 ("early intervention" or "early detection" or "early support").ti. 4739
22 or/17-21 39922
23 forensic psychiatry/ or forensic psychology/ or hospital patient/ or hospitalized adolescent/ or hospitalized child/ or hospitalized infant/ 10486
24 (forensic or inpatient* or compulsor* detention or compulsor* detained or prison* or jail*).ti,id. 55857
25 23 or 24 57926
26 22 not 25 39267
27 (systematic or structured or evidence).ti. and ((review or overview or look or examination or update* or summary).ti. or review.pt.) 43918
28 meta-analysis.pt. or (meta-analys* or meta analys* or metaanalys* or meta synth* or meta-synth* or metasynth*).ti,ab,id,hw. 53491
29 ((systematic or meta) adj2 (analys* or review)).ti,id. or ((systematic* or quantitativ* or methodologic* or qualitativ*) adj5 (review* or overview*)).ti,ab,id,sh. or ((quantitativ$ or qualitativ$) adj5 synthesis$).ti,ab,id,hw. 89819
30 (evidence adj3 review*).ti,ab,id. or "scoping review?".ti,id. or (realist adj3 review*).ti,ab,id. or (implementation adj3 review*).ti,ab,id. 26551
31 27 or 28 or 29 or 30 125296
32 26 and 31 1664
33 limit 32 to yr="2019 -Current" 737
34 limit 33 to "remove medline records" 382

**Cochrane**

#1 MeSH descriptor: [Anxiety Disorders] explode all trees 10189
#2 MeSH descriptor: [Dissociative Disorders] explode all trees 180
#3 MeSH descriptor: [Feeding and Eating Disorders] explode all trees 2486
#4 MeSH descriptor: [Mood Disorders] explode all trees 20181
#5 MeSH descriptor: [Personality Disorders] explode all trees 1838
#6 MeSH descriptor: [Schizophrenia Spectrum and Other Psychotic Disorders] explode all trees 12240
#7 MeSH descriptor: [Trauma and Stressor Related Disorders] explode all trees 4558
#8 MeSH descriptor: [Mental Health] explode all trees 3235

#9 (mood disorder* or affective disorder* or depression or depressive or dysthymi* or anxiety disorder* or agoraphobia or obsess* or compulsi* or panic or phobi* or ptsd or posttrauma* or post trauma* or affective symptoms or personality disorder* or psychos#s or psychotic or schizophrenia or eating disorder* or anorexia or bulimia):ti,kw 119235
#10 ((mental* or psyc*) near/2 (health or well* or condition* or distress or disorder or ill* or problem* or disease* or issue*)):ti,ab,kw 70133
#11 ((“at risk mental state” or “at-risk mental state” or prodrom*)):ti,ab,kw 1070
#12 {OR #1-#11} 165835
#13 pathway* near/3 (care or mental or psyc* or service* or model or referral* or help or contact*):ti,ab,kw 1666
#14 ((Referral* or treatment or "mental health service*") near/2 (pattern* or delay)):ti,ab,kw 1903
#15 (contact near/3 (service* or professional*)):ti,ab,kw 513
#16 ((helpseek* or help-seek* or help seek*) near/3 (contact* or experienc* or step* or delay or duration)):ti,ab,kw 771
#17 ("improv* access" or "point* of entry" or "journey of care" or (entry near/2 (care or service* or treatment))):ti,ab,kw 651
#18 ("early intervention" or "early detection" or ((integrat* or multidisciplinary or multi-disciplinary or coordinat* or co-ordinat* or complex) adj2 (care or treatment or therapy or team* or service or approach or intervention)) or "early support" or "support hub*" or "preventative mental health"):ti,ab,kw10912
#19 ((fast or faster or early) adj2 (access or referral* or signpost* or support or "mental health" or treatment)):ti,ab,kw0
#20 (indicated adj (prevention? or intervention? or support or help or care or approach or approaches or service? or program*)):ti,kw 0
#21 {OR #13-#20} 16203
#22 (systematic or structured or evidence) and ((review or overview or update* or summary) or review) OR meta-analysis or (meta-analys* or meta analys* or metaanalys* or meta synth* or meta-synth* or metasynth*) 221040
#23 ((systematic or meta) near/2 (analys* or review)) or ((systematic* or quantitativ* or methodologic* or qualitativ*) near/5 (review* or overview*)) or (quantitativ$ or qualitativ$ near/5 synthesis$) 40380
#24 (scoping NEXT review?) or (evidence near/3 review*) or (realist near/3 review*) or (implementation near/3 review*) 10206
#25 {OR #22-#24} 221598
#26 #12 and #21 and #25 with Cochrane Library publication date Between Jan 2019 and Apr 2024, in Cochrane Reviews (Word variations have been searched) 15

### Appendix 3: AMSTAR2 items, adaptations and ratings

AMSTAR2 item descriptions

1. Did the research questions and inclusion criteria for the review include the components of PICO?
2. Did the report of the review contain an explicit statement that the review methods were established prior to the conduct of the review and did the report justify any significant deviations from the protocol?
3. Did the review authors explain their selection of the study designs for inclusion in the review?
4. Did the review authors use a comprehensive literature search strategy?
5. Did the review authors perform study selection in duplicate?
6. Did the review authors perform data extraction in duplicate?
7. Did the review authors provide a list of excluded studies and justify the exclusions?
8. Did the review authors describe the included studies in adequate detail?
9. Did the review authors use a satisfactory technique for assessing the risk of bias (RoB) in individual studies that were included in the review?
10. Did the review authors report on the sources of funding for the studies included in the review?
11. If meta-analysis was performed, did the review authors use appropriate methods for statistical combination of results?
12. If meta-analysis was performed, did the review authors assess the potential impact of RoB in individual studies on the results of the meta-analysis or other evidence synthesis?
13. Did the review authors account for RoB in primary studies when interpreting/discussing the results of the review?
14. Did the review authors provide a satisfactory explanation for, and discussion of, any heterogeneity observed in the results of the review?
15. If they performed quantitative synthesis did the review authors carry out an adequate investigation of publication bias (small study bias) and discuss its likely impact on the results of the review?
16. Did the review authors report any potential sources of conflict of interest, including any funding they received for conducting the review?

In the AMSTAR 2 tool there are 7 critical domains and 6 non-critical domains. If a review had no critical flaws and none/one non-critical weakness, it was deemed as high quality. Reviews with no critical flaws but one or more non-critical weakness were rated as moderate quality. Reviews with one critical flaw (irrespective of non-critical weaknesses) were judged as low quality, and those with more than one critical flaw (irrespective of non-critical) were rated as critically low quality. According to AMSTAR 2 guidelines, reviews assessed as critically low in quality should not be relied upon for providing accurate and dependable literature summaries (Shea et al., 2017).

Adaptations to AMSTAR2 criteria for scoping reviews and qualitative meta-syntheses
Adaptations according to Cooper et al. (2024) review:
Item 1: For Yes – removed the requirement of “comparator group” in research question and inclusion criteria.

Item 2: For Partial yes – removed the requirement of inclusion of “risk of bias assessment” in the protocol. For yes – removed the requirement of inclusion of “plan for investigating causes of heterogeneity” in protocol (scoping and qualitative reviews only).
Item 8: For Partial yes – removed the requirement of “comparators” in description of studies.
Item 9: Marked this question as NA (scoping and qualitative reviews only).
Item 13: Marked as NA for scoping review and qualitative syntheses (in line with 2).
Items 11,12 and 15 were automatically marked as NA for scoping review and qualitative studies, as well as those not reporting meta-analyses.

Re-classifying critical domain:
In the rating of reviews using “critical” and “non-critical weaknesses” identified by Shea et al. (2017), the domains “*Did the review authors provide a list of excluded studies and justify the exclusions?*” (item 7) was originally deemed critical. However, after discussion with the research team it was decided that given common practice in this field it could be considered *non-critical*. We therefore considered items 2, 4, 9, 11, 13 and 15 (when not marked as NA) to be critical. In the following table, ratings in **<u>bold underline</u>** signify critical weaknesses resulting in low or critically low-quality ratings.

In the Results section we report the quality appraisal of studies using the amended metrics.

**AMSTAR2 ratings for each study (items 1-16 listed above)**

| **AMSTAR2 criteria** | **1** | **2** | **3** | **4** | **5** | **6** | **7** | **8** | **9 RCT** | **9 NRSI** | **10** | **11 RCT** | **11 NRSI** | **12** | **13** | **14** | **15** | **16** | **Overall Score** |
|---|---|---|---|---|---|---|---|---|---|---|---|---|---|---|---|---|---|---|---|
| **Aceituno 2021** | Yes | Yes | Yes | Partial yes | Yes | Yes | Partial yes | Partial yes | N/A | N/A | No | N/A | N/A | N/A | N/A | No | N/A | Yes | Moderate |
| **Causier 2024** | Yes | Yes | Yes | Partial yes | Yes | Yes | Partial yes | Partial yes | N/A | N/A | No | N/A | N/A | N/A | N/A | Yes | N/A | Yes | High |

| | | | | | | | | | | | | | | | | | | | |
|---|---|---|---|---|---|---|---|---|---|---|---|---|---|---|---|---|---|---|---|
| **Farooq 2024** | Yes | Yes | Yes | Yes | Yes | Yes | Partial yes | Yes | Yes | Yes | No | N/A | N/A | N/A | Yes | No | N/A | Yes | Moderate |
| **Hamson 2023** | Yes | Yes | No | Yes | Yes | Yes | Yes | Yes | Yes | Yes | Yes | N/A | N/A | N/A | Yes | No | N/A | Yes | Moderate |
| **Jongen 2023** | Yes | Yes | Yes | Partial yes | Yes | Yes | No | Partial yes | N/A | N/A | No | N/A | N/A | N/A | N/A | No | N/A | Yes | Moderate |
| **Koreshe 2023** | Yes | **No** | Yes | Yes | Yes | Yes | No | Yes | N/A | N/A | No | N/A | N/A | N/A | **No** | No | N/A | Yes | Critically Low |
| **Loughlin 2020** | Yes | **No** | Yes | Partial yes | No | No | No | Partial yes | N/A | N/A | No | N/A | N/A | N/A | N/A | No | N/A | No | Low |

| | | | | | | | | | | | | | | | | | | | |
|---|---|---|---|---|---|---|---|---|---|---|---|---|---|---|---|---|---|---|---|
| **Murden 2024** | Yes | Partial yes | Yes | Partial yes | Yes | Yes | No | Yes | N/A | Yes | No | N/A | N/A | N/A | **No** | No | N/A | Yes | Low |
| **O'Connell 2021** | Yes | Yes | No | **No** | Yes | No | No | No | N/A | N/A | No | N/A | N/A | N/A | N/A | No | N/A | Yes | Low |
| **Pehlivan 2022** | Yes | **No** | Yes | Partial yes | Yes | No | No | Partial yes | **No** | **No** | No | N/A | N/A | N/A | **No** | No | N/A | Yes | Critically Low |
| **Puntis 2020** | Yes | Yes | Yes | Yes | Yes | Yes | Yes | Yes | Yes | N/A | Yes | Yes | N/A | Yes | Yes | Yes | Yes | Yes | High |
| **Ratheesh 2022** | Yes | Partial yes | No | Yes | Yes | Yes | No | Partial yes | Yes | Yes | No | N/A | N/A | N/A | Yes | Yes | N/A | Yes | Moderate |

| | | | | | | | | | | | | | | | | | | | |
|---|---|---|---|---|---|---|---|---|---|---|---|---|---|---|---|---|---|---|---|
| **Salazar de Pablo 2024** | Yes | Partial yes | No | Partial yes | Yes | Yes | No | Partial yes | Partial yes | Yes | No | Yes | Yes | N/A | Yes | Yes | Yes | Yes | Moderate |
| **Settipani 2019** | Yes | Yes | Yes | Yes | Yes | Yes | No | Yes | N/A | N/A | No | N/A | N/A | N/A | N/A | No | N/A | Yes | Moderate |
| **Tiller 2023** | Yes | Yes | No | <u>**No**</u> | Yes | No | No | No | N/A | N/A | No | N/A | N/A | N/A | N/A | No | N/A | Yes | Low |
| **Williams 2024** | Yes | Yes | Yes | Yes | Yes | Yes | No | Yes | Yes | N/A | No | Yes | N/A | Yes | Yes | Yes | Yes | Yes | Moderate |

### Appendix 4: Excluded studies with reasons for exclusion

| Authors | Reasons for exclusion |
|---|---|
| Adler, A. J., Drown, L., Boudreaux, C., Coates, M. M., Marx, A., Akala, O., ... & Bukhman, G. (2023). Understanding integrated service delivery: a scoping review of models for noncommunicable disease and mental health interventions in low-and-middle income countries. BMC health services research, 23(1), 99. | Wrong diagnosis |
| Adu, J., Oudshoorn, A., Van Berkum, A., Pervez, R., Norman, R., Canas, E., ... & MacDougall, A. G. (2022). System transformation to enhance transitional age youth mental health–a scoping review. Child and adolescent mental health, 27(4), 399-418. | Not early intervention |
| Albert, N., & Weibell, M. A. (2019). The outcome of early intervention in first episode psychosis. International Review of Psychiatry, 31(5-6), 413-424. | Wrong study design |
| Allan, S. M., Hodgekins, J., Beazley, P., & Oduola, S. (2021). Pathways to care in at-risk mental states: A systematic review. Early intervention in Psychiatry, 15(5), 1092-1103. | Not early intervention |
| Baudinet, J., & Simic, M. (2021). Adolescent eating disorder day programme treatment models and outcomes: a systematic scoping review. Frontiers in psychiatry, 12, 652604. | Not early intervention |
| Breneol, S., Doucet, S., McIsaac, J. L., Riveroll, A., Cassidy, C., Charlton, P., ... & Curran, J. A. (2022). Programmes to support transitions in community care for children with complex care needs: a scoping review. BMJ open, | Wrong diagnosis |
| Bryant, E., Spielman, K., Le, A., Marks, P., Touyz, S., & Maguire, S. (2022). Screening, assessment and diagnosis in the eating disorders: findings from a rapid review. Journal of Eating Disorders, 10(1), 78. | No intervention complexity |
| Campaña, I. C. J., Romero-Galisteo, R. P., Manzanares, M. T. L., & Morales, N. M. (2019). Evaluation of quality of service in Early Intervention: A systematic review. Anales de Pediatría (English Edition), 90(5), 301-309. | Wrong outcome |
| Carrà, G., Bartoli, F., Capogrosso, C. A., Cioni, R. M., Moretti, F., Piacenti, S., ... & Bebbington, P. E. (2022). Innovations in community-based mental health care: an overview of meta-analyses. International Review of Psychiatry, 34(7-8), 770-782. | Wrong study design |
| Carrier, J. D., Roberge, P., Vanasse, A., Gallagher, F., Paradis-Gagné, E., Maillet, L., & Jacques, M. C. (2023). Modélisation des cibles potentielles d'amélioration de l'accès aux interventions cognitivo-comportementales pour les personnes avec troubles anxieux (Doctoral dissertation, Université de Sherbrooke). | No intervention complexity |
| Chan, S. K., Chan, H. Y., Devlin, J., Bastiampillai, T., Mohan, T., Hui, C. L., ... & Chen, E. Y. (2019). A systematic review of long-term outcomes of patients with psychosis who received early intervention services. International Review of Psychiatry, 31(5-6), 425-440. | Fewer than 3 databases / no quality appraisal for systematic review |
| Chetty, A., Guse, T., & Malema, M. (2023). Integrated vs non-integrated treatment outcomes in dual diagnosis disorders: A systematic review. Health SA Gesondheid, 28, 2094. | Not early intervention |

| | |
|---|---|
| Coates, D., Coppleson, D., & Schmied, V. (2020). Integrated physical and mental healthcare: an overview of models and their evaluation findings. JBI Evidence Implementation, 18(1), 38-57. | Special population |
| Cooper, M., Avery, L., Scott, J., Ashley, K., Jordan, C., Errington, L., & Flynn, D. (2022). Effectiveness and active ingredients of social prescribing interventions targeting mental health: a systematic review. BMJ open, 12(7), e060214 | No intervention complexity |
| Cosh, S. M., McNeil, D. G., Jeffreys, A., Clark, L., & Tully, P. J. (2023). Athlete mental health help-seeking: A Systematic review and meta-analysis of rates, barriers and facilitators. Psychology of Sport and Exercise, 102586. | Special population |
| Curtis J.; McHugh C.; Hodgins M.; Hu N.; Eapen V.; Lingam R. A framework for evaluating the implementation and maintenance of integration within the youth mental health system. Early Intervention in Psychiatry 17(S1) 323 https://dx.doi.org/10.1111/eip.13409 | Wrong study design |
| De Giuseppe, R., Di Napoli, I., Porri, D., & Cena, H. (2019). Pediatric obesity and eating disorders symptoms: the role of the multidisciplinary treatment. A systematic review. Frontiers in pediatrics, 7, 123. | Wrong diagnosis |
| de Oliveira, J. L., Dal Sasso Mendes, K., de Almeida, L. Y., de Almeida, J. C. P., Souza Gonçalves, J., Strobbe, S., & de Souza, J. (2023). Mental Health Care in Primary Health Care: An Integrative Review. Issues in Mental Health Nursing, 44(4), 329-337. | Not early intervention |
| Devoe, D. J., Farris, M. S., Townes, P., & Addington, J. (2019). Attenuated psychotic symptom interventions in youth at risk of psychosis: A systematic review and meta-analysis. Early intervention in psychiatry, 13(1), 3-17. | Not early intervention |
| Devoe, D., & Addington, J. (2019). T29. Treatment and global functioning in youth at clinical high risk for psychosis: a systematic review and meta-analysis. Schizophrenia Bulletin, 45(Suppl 2), S214. | Wrong study design |
| Eghaneyan, B. H., & Murphy, E. R. (2019). Mental health help-seeking experiences of Hispanic women in the United States: results from a qualitative interpretive meta-synthesis. Social Work in Public Health, 34(6), 505-518 | Not early intervention |
| Erzin, G., & Gülöksüz, S. (2021). Early interventions for clinical high-risk state for psychosis. Archives of Neuropsychiatry, 58(Suppl 1), S7. | Wrong study design |
| Esponda, G. M., Hartman, S., Qureshi, O., Sadler, E., Cohen, A., & Kakuma, R. (2020). Barriers and facilitators of mental health programmes in primary care in low-income and middle-income countries. The Lancet Psychiatry, [illegible] | Not early intervention |
| Estrade, A., Salazar de Pablo, G., Zanotti, A., Wood, S., Fisher, H. L., & Fusar-Poli, P. (2022). Public health primary prevention implemented by clinical high-risk services for psychosis. Translational Psychiatry, 12(1), 43. | Not early intervention |
| Fang, M., Fan, Z., Liu, S., Feng, S., Zhu, H., Yin, D., ... & Wang, G. (2023). Preventive interventions for individuals at risk of developing bipolar disorder: A systematic review and meta-analysis. Journal of Affective Disorders | No intervention complexity |
| Fantuzzi, C., & Mezzina, R. (2020). Dual diagnosis: A systematic review of the organization of community health services. International Journal of Social Psychiatry, 66(3), 300-310. | Not early intervention |

| | |
|---|---|
| Filia, K., Eastwood, O., Herniman, S., & Badcock, P. (2021). Facilitating improvements in young people's social relationships to prevent or treat depression: A review of empirically supported interventions. Translational Psychiatry, 11(1), 305. | Wrong study design |
| Fisak, B., Griffin, K., Nelson, C., Gallegos-Guajardo, J., & Davila, S. (2023). The effectiveness of the FRIENDS programs for children and adolescents: a meta-analytic review. Mental Health & Prevention, 30, 200271. | No intervention complexity |
| Fusar-Poli, P. (2019). Integrated mental health services for the developmental period (0 to 25 years): a critical review of the evidence. Frontiers in Psychiatry, 10, 355. | Wrong study design |
| Gaebel, W., Kerst, A., Janssen, B., Becker, T., Musalek, M., Rössler, W., ... & Stricker, J. (2020). EPA guidance on the quality of mental health services: A systematic meta-review and update of recommendations focusing on care coordination. European Psychiatry, 63(1), e75. | Wrong study design |
| Gee, B., Reynolds, S., Carroll, B., Orchard, F., Clarke, T., Martin, D., ... & Pass, L. (2020). Practitioner Review: Effectiveness of indicated school-based interventions for adolescent depression and anxiety–a meta-analytic review. Journal of Child Psychology and Psychiatry, 61(7), 739-756. | No intervention complexity |
| Gee, B., Wilson, J., Clarke, T., Farthing, S., Carroll, B., Jackson, C., ... & Notley, C. (2021). Delivering mental health support within schools and colleges–a thematic synthesis of barriers and facilitators to implementation of indicated psychological interventions for adolescents. Child and adolescent mental health, 26(1), 34-46. | No intervention complexity |
| Gerstl, B., Ahinkorah, B. O., Nguyen, T. P., John, J. R., Hawker, P., Winata, T., ... & Eapen, V. (2024). Evidence-based long term interventions targeting acute mental health presentations for children and adolescents: systematic review. Frontiers in Psychiatry, 15, 1324220. V. | Not early intervention |
| Gimba, S. M., Harris, P., Saito, A., Udah, H., Martin, A., & Wheeler, A. J. (2020). The modules of mental health programs implemented in schools in low-and middle-income countries: findings from a systematic literature review. BMC public health, 20, 1-10. | No intervention complexity |
| Glover-Wright, C., Coupe, K., Campbell, A. C., Keen, C., Lawrence, P., Kinner, S. A., & Young, J. T. (2023). Health outcomes and service use patterns associated with co-located outpatient mental health care and alcohol and other drug specialist treatment: A systematic review. Drug and | Not early intervention |
| Grunze, H., Schaefer, M., Scherk, H., Born, C., & Preuss, U. W. (2021). Comorbid bipolar and alcohol use disorder—a therapeutic challenge. Frontiers in psychiatry, 12, 660432. | Wrong study design |
| Gunawardena, H., Voukelatos, A., Nair, S., Cross, S., & Hickie, I. B. (2023). Efficacy and effectiveness of universal school-based wellbeing interventions in Australia: A systematic review. International Journal of Environmental Research and Public Health, 20(15), 6508. | Not early intervention |
| Harvey, C., Zirnsak, T. M., Brasier, C., Ennals, P., Fletcher, J., Hamilton, B., ... & Brophy, L. (2023). Community-based models of care facilitating the recovery of people living with persistent and complex mental health needs: [illegible] | Wrong diagnosis |

| | |
|---|---|
| Hazell, P. (2021). Updates in treatment of depression in children and adolescents. Current opinion in psychiatry, 34(6), 593-599. | Wrong study design |
| Hugh-Jones, S., Beckett, S., Tumelty, E., & Mallikarjun, P. (2021). Indicated prevention interventions for anxiety in children and adolescents: a review and meta-analysis of school-based programs. European Child & Adolescent Psychiatry, 30(6), 849-860 | No intervention complexity |
| Hui, T. T., Garvey, L., & Olasoji, M. (2021). Improving the physical health of young people with early psychosis with lifestyle interventions: Scoping review. International Journal of Mental Health Nursing, 30(6), 1498-1524. | Wrong outcomes |
| Isaacs, A. N., & Mitchell, E. K. (2024). Mental health integrated care models in primary care and factors that contribute to their effective implementation: a scoping review. International Journal of Mental Health | Not early intervention |
| Jeindl, R., Hofer, V., Bachmann, C., & Zechmeister-Koss, I. (2023). Optimising child and adolescent mental health care–a scoping review of international best-practice strategies and service models. Child and Adolescent Psychiatry and Mental Health, 17(1), 135. | Not early intervention |
| Kalindjian, N., Hirot, F., Stona, A. C., Huas, C., & Godart, N. (2021). Early detection of eating disorders: a scoping review. Eating and Weight Disorders-Studies on Anorexia, Bulimia and Obesity, 1-48. | No intervention complexity |
| Karapareddy, V. (2019). A review of integrated care for concurrent disorders: cost effectiveness and clinical outcomes. Journal of Dual Diagnosis, 15(1), 56-66. | Wrong outcomes |
| Kerbage, H., Bazzi, O., El Hage, W., Corruble, E., & Purper-Ouakil, D. (2022, April). Early interventions to prevent post-traumatic stress disorder in youth after exposure to a potentially traumatic event: A scoping review. In Healthcare (Vol. 10, No. 5, p. 818). MDPI. | No intervention complexity |
| Krishnamoorthy, G., Shin, S. M., & Rees, B. (2023). Day Programs for children and adolescents with eating disorders: A systematic review. European Eating Disorders Review, 31(2), 199-225. | Not early intervention |
| Lenton-Brym, T., Rodrigues, A., Johnson, N., Couturier, J., & Toulany, A. (2020). A scoping review of the role of primary care providers and primary care-based interventions in the treatment of pediatric eating disorders. Eating Disorders, 28(1), 47-66. | Not early intervention |
| Lopez-Carmen, V., McCalman, J., Benveniste, T., Askew, D., Spurling, G., Langham, E., & Bainbridge, R. (2019). Working together to improve the mental health of indigenous children: A systematic review. Children and Youth Services Review, 104, 104408. | Not early intervention |
| Lovén Wickman, U., & Schmidt, M. (2023). Experiences of primary care among young adults with mental illness–A systematic literature review. Scandinavian Journal of Caring Sciences, 37(3), 628-641. | Not early intervention |
| Lynch, L., Moorhead, A., Long, M., & Hawthorne-Steele, I. (2023). The role of Informal sources of help in Young people's Access to, Engagement with, and maintenance in Professional Mental Health Care—A Scoping Review. Journal of Child and Family Studies, 32(11), 3350-3365. | No intervention complexity |
| Ma, S., Yu, H., Liang, N., Zhu, S., Li, X., Robinson, N., & Liu, J. (2020). Components of complex interventions for healthcare: A narrative synthesis of qualitative studies. Journal of Traditional Chinese Medical Sciences | Wrong diagnosis |

| | |
|---|---|
| MacDonald, K., Ferrari, M., Fainman-Adelman, N., & Iyer, S. N. (2021). Experiences of pathways to mental health services for young people and their carers: a qualitative meta-synthesis review. Social psychiatry and psychiatric epidemiology, 56, 339-361. | Not early intervention |
| Mascayano, F., van der Ven, E., Martinez-Ales, G., Henao, A. R., Zambrano, J., Jones, N., ... & Dixon, L. B. (2021). Disengagement from early intervention services for psychosis: a systematic review. Psychiatric Services, 72(1), 49-60. | Wrong outcomes |
| McDermott, E., Eastham, R., Hughes, E., Pattinson, E., Johnson, K., Davis, S., ... & Jenzen, O. (2021). Explaining effective mental health support for LGBTQ+ youth: A meta-narrative review. SSM-mental health, 1, 100004. | Wrong outcomes |
| McHugh C.; Curtis J. (2023). Integrated care models for youth mental health: A systematic review and meta-analysis. Early Intervention in Psychiatry 17( S1) 204–332 https://doi.org/10.1111/eip.13409 | Wrong study design |
| McKenna, J. W., Brigham, F., Garwood, J., Zurawski, L., Koc, M., Lavin, C., & Werunga, R. (2021). A systematic review of intervention studies for young children with emotional and behavioral disorders: identifying the research base. Journal of Research in Special Educational Needs, 21(2), 120-145. | Wrong diagnosis |
| Michelson, D., Hodgson, E., Bernstein, A., Chorpita, B. F., & Patel, V. (2022). Problem solving as an active ingredient in indicated prevention and treatment of youth depression and anxiety: An integrative review. Journal of Adolescent Health, 71(4), 390-405. | No intervention complexity |
| Miller, E., Bosun-Arjie, S. F., & Ekpenyong, M. S. (2021). Black and ethnic minority carers perceptions on mental health services and support in the United Kingdom: a systematic review. Journal of Public Mental Health, 20(4), 298-311. | Wrong outcomes |
| Murphy, R., Huggard, L., Fitzgerald, A., Hennessy, E., & Booth, A. (2024). A systematic scoping review of peer support interventions in integrated primary youth mental health care. Journal of Community Psychology, 52(1), 154-180 | No intervention complexity |
| Nicula, M., Pellegrini, D., Grennan, L., Bhatnagar, N., McVey, G., & Couturier, J. (2022). Help-seeking attitudes and behaviours among youth with eating disorders: a scoping. | Not early interventions |
| Nigatu, Y. T., Huang, J., Rao, S., Gillis, K., Merali, Z., & Wang, J. (2019). Indicated prevention interventions in the workplace for depressive symptoms: a systematic review and meta-analysis. American journal of preventive medicine, 56(1), e23-e33. | Special population |
| Olson, J. R., Benjamin, P. H., Azman, A. A., Kellogg, M. A., Pullmann, M. D., Suter, J. C., & Bruns, E. J. (2021). Systematic review and meta-analysis: Effectiveness of wraparound care coordination for children and adolescents. Journal of the American Academy of Child & Adolescent Psychiatry, 60(11), 1353-1366. | Not early intervention |
| Oluwoye, O., Davis, B., Kuhney, F. S., & Anglin, D. M. (2021). Systematic review of pathways to care in the US for Black individuals with early psychosis. npj Schizophrenia, 7(1), 58. | Not early intervention |
| Oluwoye, O., Nagendra, A., Kriegel, L. S., Anglin, D. M., Santos, M. M., & López, S. R. (2023). Reorienting the focus from an individual to a community-level lens to improve the pathways through care for early psychosis in the United States. SSM-Mental Health, 3, 100209. | Wrong study design |

| | |
|---|---|
| Oosterbaan, V., Covers, M. L., Bicanic, I. A., Huntjens, R. J., & de Jongh, A. (2019). Do early interventions prevent PTSD? A systematic review and meta-analysis of the safety and efficacy of early interventions after sexual assault. European journal of psychotraumatology, 10(1), 1682932. | No intervention complexity |
| Page, I. S., Leitch, E., Gossip, K., Charlson, F., Comben, C., & Diminic, S. (2022). Modelling mental health service needs of Aboriginal and Torres Strait Islander peoples: a review of existing evidence and expert consensus. Australian and New Zealand Journal of Public Health, 46(2), 177-185. | Wrong outcome |
| Pipkin, A. (2021). Evidence base for early intervention in psychosis services in rural areas: A critical review. Early Intervention in Psychiatry, 15(4), 762-774. | Wrong study design |
| Poletti, M. (2022). Early intervention services for youth at clinical high-risk for psychosis: the Reggio Emilia at-risk mental state (ReARMS) experience. Rivista sperimentale di freniatria: la rivista dei servizi di salute mentale: CXLVI, 3, 2022, 61-80. | Wrong study design |
| Radez, J., Waite, F., Izon, E., & Johns, L. (2023). Identifying individuals at risk of developing psychosis: A systematic review of the literature in primary care services. Early Intervention in Psychiatry, 17(5), 429-446. | Wrong outcomes |
| Radunz, M., Ali, K., & Wade, T. D. (2023). Pathways to improve early intervention for eating disorders: Findings from a systematic review and meta-analysis. International Journal of Eating Disorders, 56(2), 314-330. | Not early intervention |
| Reinares, M., Martínez-Arán, A., & Vieta, E. (Eds.). (2019). Psychotherapy for bipolar disorders: An integrative approach. | Wrong study design |
| Richardson, A., Richard, L., Gunter, K., Cunningham, R., Hamer, H., Lockett, H., ... & Derrett, S. (2020). A systematic scoping review of interventions to integrate physical and mental healthcare for people with serious mental illness and substance use disorders. Journal of Psychiatric Research, 128, 52-67. | Special population |
| Roberts NP, Kitchiner NJ, Kenardy J, Robertson L, Lewis C, Bisson JI. Multiple session early psychological interventions for the prevention of post-traumatic stress disorder. Cochrane Database of Systematic Reviews 2019, Issue 8. Art. No.: CD006869. DOI: 10.1002/14651858.CD006869.pub3. Accessed 25 October 2024. | No intervention complexity |
| Robson, E., & Greenwood, K. (2022). Rates and predictors of disengagement and strength of engagement for people with a first episode of psychosis using early intervention services: a systematic review of predictors and meta-analysis of disengagement rates. Schizophrenia Bulletin Open, 3(1), sgac012. | Wrong outcomes |
| Rosic, T., Lovell, E., Macmillan, H., Samaan, Z., & Morgan, R. L. (2023). Components of Outpatient Child and Youth Concurrent Disorders Programs: A Critical Interpretive Synthesis. Canadian Journal of psychiatry. Revue Canadienne de Psychiatrie, 7067437231212037-7067437231212037. | Fewer than 3 databases / no quality appraisal for systematic review |
| Sangsawang, B., Wacharasin, C., & Sangsawang, N. (2019). Interventions for the prevention of postpartum depression in adolescent mothers: a systematic review. Archives of women's mental health, 22, 215-228. | Not early intervention |

| | |
|---|---|
| Saraf, G., Moazen-Zadeh, E., Pinto, J. V., Ziafat, K., Torres, I. J., Kesavan, M., & Yatham, L. N. (2021). Early intervention for people at high risk of developing bipolar disorder: a systematic review of clinical trials. The Lancet Psychiatry, 8(1), 64-75. | No intervention complexity |
| Saraf, G., Moazen-Zadeh, E., Pinto, J. V., Ziafat, K., Torres, I. J., Kesavan, M., & Yatham, L. N. (2021). Early intervention for people at high risk of developing bipolar disorder: a systematic review of clinical trials. The Lancet Psychiatry, 8(1), 64-75 | Wrong study design |
| Sarakbi, D., Groll, D., Tranmer, J., & Sears, K. (2022). Achieving Quality Integrated Care for Adolescent Depression: A Scoping Review. Journal of Primary Care & Community Health, 13, 21501319221131684. | Not early intervention |
| Sheridan Rains, L., Echave, A., Rees, J., Scott, H. R., Lever Taylor, B., Broeckelmann, E., ... & Johnson, S. (2021). Service user experiences of community services for complex emotional needs: A qualitative thematic synthesis. PLoS One, 16(4), e0248316. | Not early intervention |
| Singh, G., Narang, I., & Marwaha, R. (2022, October). Integrated Care in Pediatric Depression: A Review. In AACAP/CACAP 2022 Annual Meeting. AACAP. | Wrong study design |
| So, M., McCord, R. F., & Kaminski, J. W. (2019). Policy levers to promote access to and utilization of children's mental health services: a systematic review. Administration and Policy in Mental Health and Mental Health Services Research, 46(3), 334-351. | Not early intervention |
| Spettigue, W. J., Obeid, N., Allan, A., Seale, R., Porter, J., & Norris, M. L. (2022, October). Early Intervention for Children and Adolescents With Eating Disorders: A Scoping Review of the Literature. In AACAP/CACAP 2022 Annual Meeting. AACAP. | Wrong study design |
| Ssegonja, R., Nystrand, C., Feldman, I., Sarkadi, A., Langenskiöld, S., & Jonsson, U. (2019). Indicated preventive interventions for depression in children and adolescents: A meta-analysis and meta-regression. Preventive medicine, 118, 7-15. | No intervention complexity |
| Stein, B., Müller, M. M., Meyer, L. K., & Söllner, W. (2020). CL Guidelines Working Group. Psychiatric and psychosomatic consultation-liaison services in general hospitals: a systematic review and meta-analysis of effects on symptoms of depression and anxiety. Psychother. Psychosom., 89, 6-16. | Wrong setting |
| Tahmazov, E., Blachier, A., Nabbe, P., Guillou-Landreat, M., Walter, M., & Lemey, C. (2024). Effect of early intervention for early-stage psychotic disorders on suicidal behaviours–a systematic review protocol. Frontiers in Psychiatry, 15, 1359764. | Wrong study design |
| Tahmazov, E., Lemey, C., & Walter, M. (2020). Suicidal behavior in early psychosis. European Psychiatry, 63. | Wrong study design |
| van Genk, C., Roeg, D., van Vugt, M., van Weeghel, J., & Van Regenmortel, T. (2023). Current insights of community mental healthcare for people with severe mental illness: A scoping review. Frontiers in Psychiatry, 14, 1156235. | Not early intervention |

| | |
|---|---|
| Waid, J., Halpin, K., & Donaldson, R. (2021). Mental health service navigation: a scoping review of programmatic features and research evidence. Social Work in Mental Health, 19(1), 60-79. | Not early intervention |
| Westby, M., Ijaz, S., Savović, J., McLeod, H., Dawson, S., Welsh, T., ... & Bradley, N. (2024). Virtual wards for people with frailty: what works, for whom, how and why—a rapid realist review. *Age and Ageing*, *53*(3), afae039. | Wrong diagnosis |
| Whitfield, J., Owens, S., Bhat, A., Felker, B., Jewell, T., & Chwastiak, L. (2023). Successful ingredients of effective Collaborative Care programs in low-and middle-income countries: A rapid review. Cambridge Prisms: Global Mental Health, 10, e11. | Not early intervention |
| Wolitzky-Taylor, K. (2023). Integrated behavioral treatments for comorbid anxiety and substance use disorders: A model for understanding integrated treatment approaches and meta-analysis to evaluate their efficacy. Drug and Alcohol Dependence, 110990. | Wrong diagnosis |
| Yonek, J., Lee, C. M., Harrison, A., Mangurian, C., & Tolou-Shams, M. (2020). Key components of effective pediatric integrated mental health care models: a systematic review. JAMA pediatrics, 174(5), 487-498. | Not early intervention |
| Zechmeister-Koss, I., Jeindl, R., & Hofer, V. (2023). OP33 Child And Adolescent Mental Health Care Models: A Scoping Review. International Journal of Technology Assessment in Health Care, 39(S1), S9-S9. | Wrong study design |
| Zhang, Y., Kwekkeboom, K., Kim, K. S., Loring, S., & Wieben, A. M. (2020). Systematic review and meta-analysis of psychosocial uncertainty management interventions. Nursing Research, 69(1), 3-12. | Wrong diagnosis |

### Appendix 5: Overlapping studies

| **Two reviews** | **Three reviews** |
|---|---|
| Valencia 2012 (1) | Bertelsen 2008 (2) |
| McCann 2011 (3) | Connor 2016 (4) |
| Bay 2016 (5) | Total: 2 studies |
| Islam 2015 (6) | |
| McClelland 2018 (7) | |
| Rosling 2016 (8) | |
| Jansen 2018 (9) | |
| Iyer 2015 (10) | |
| Craig 2004 (11) | |

| |
|---|
| Kane 2016 (12) |
| Gafoor 2010 (13) |
| Grawe 2006 (14) |
| Srihari 2015 (15) |
| Wade 2017 (16) |
| Watson 2016 (17) |
| Lloyd-Evans 2015 (18) |
| Chan 2018 (19) |
| Chong 2005 (20) |
| McGorry 1996 (21) |
| Krstev 2004 (22) |
| Malla 2005 (23) |
| Malla 2014 (24) |
| Cassidy 2008 (25) |
| Melle 2004 (26) |
| Joa 2008 (27) |
| Ferrara 2019 (28) |
| Total: 26 studies |
